# Towards connectome-guided optimization of deep brain stimulation for gait dysfunction

**DOI:** 10.64898/2026.03.21.26348543

**Authors:** Calvin W. Howard, Savir Madan, Lan Luo, Nanditha Rajamani, Lukas L. Goede, Lauren A. Hart, Ella Gray Settle, Martin Reich, Andreas Horn, Michael D. Fox

## Abstract

Despite the success of deep brain stimulation (DBS) in treating many symptoms of Parkinson’s Disease (PD), treatment of gait dysfunction has remained challenging. Recent work suggests that gait dysfunction may require activation of different electrode contacts connected to different brain circuits than those used to treat other PD symptoms. In this study we developed an algorithm to select DBS contacts and parameters based on the location of a patient’s implanted electrodes that maximizes overlap of the modeled stimulation volume with brain circuitry associated with gait dysfunction. We trained and tested this algorithm using two independent cohorts of Parkinson’s STN DBS patients who had previously undergone clinical optimization of their DBS settings (training n = 44; test n = 100). We found that the ‘gait-optimized’ stimulation volumes differed greatly from the patients’ clinically selected stimulation volumes, with use of different electrode contacts in over 85% of patients. Increased similarity between the ‘gait-optimized’ stimulation volume and a patient’s clinical stimulation volume was correlated with gait improvement after DBS. To illustrate how this information might be used clinically, we reprogrammed six PD patients from their clinical settings to the ‘gait-optimized’ settings, and found all patients reported subjective gait improvement. These findings suggest that the optimal DBS settings for gait differ from clinically selected DBS settings and that a connectome-based algorithm might help guide DBS reprogramming to improve gait function.

## Introduction

Despite efficacy of deep brain stimulation (DBS) for many symptoms of Parkinson disease, treatment of gait dysfunction with DBS remains challenging.^1–4^ While there are many potential explanations,^5–11^ one possibility is that the optimal stimulation site for gait dysfunction differs from the sites commonly used to treat tremor, rigidity, or bradykinesia.^7,12^ Clinical practice often focuses on these three symptoms, partially because they may respond rapidly to DBS and are easily assessed for changes. In contrast, gait symptoms may not respond immediately, and often require more time-intensive assessment for changes, making programming DBS for gait dysfunction challenging.

Multiple studies have explored associations between DBS stimulation site and gait outcomes,^7,12–14^ with two recent studies having identified brain circuits correlated with DBS induced changes in gait, one to a functionally connected brain network^15^ and the other to a set of tractography fibers.^16^ It is possible that these gait-specific targets might be used to guide DBS parameters for gait dysfunction.^17^ However, it remains unclear if these gait-specific targets yield unique DBS settings or if these new settings are associated with gait benefit.

To investigate the above questions, we developed an optimization algorithm to maximize DBS stimulation volume overlap with both functional networks and fiber targets. We applied this algorithm to both the brain network and fiber tract associated with gait dysfunction to generate ‘gait-optimized’ DBS parameters.^15,16^ We investigated if the new ‘gait-optimized’ parameters differed meaningfully from standard clinical parameters by comparing their stimulation volumes. We also tested whether incidental similarity between clinical and ‘gait-optimized’ stimulation volumes was associated with gait outcomes, and lastly investigated the feasibility of reprogramming DBS to ‘gait-optimized’ settings in six patients.

## Results

### Cohort characteristics

We analyzed retrospective data from 144 patients across 2 independent Parkinson disease subthalamic nucleus (STN) DBS cohorts, and then studied 6 prospective patients with STN and globus pallidus internus (GPi) DBS (**Table 1**). The training cohort was from Würzburg, Germany (n = 44), and was used to select model hyperparameters. However, this cohort was one of the cohorts used to derive the network and fibers associated with gait dysfunction,^15,16^ and so we collected an unseen testing cohort to evaluate the algorithm without circularity. The testing cohort was from Brigham and Women’s Hospital, Boston (n = 100), and was used for evaluations throughout the manuscript. The prospective patients were patients with self-identified gait dysfunction and DBS who presented to either Beth Israel Deaconess Medical Center or Brigham and Women’s Hospital, Boston (n = 6).

**Table 1:** Overview of the Parkinson Disease DBS cohorts. Data are shown as mean ± standard deviation. Age is in years. UPDRS-III Baseline Score represents motor symptom severity prior to surgery, and UPDRS-III Score Improvement is expressed as the percent change from baseline at one year postoperatively in the ON medication/ON stimulation state.

| DBS Center | Cohort | N (#F) | Target | Age | UPDRS-III Baseline Score | UPDRS-III Score Improvement (%) | Medication State | Disease Duration | Reference |
| --- | --- | --- | --- | --- | --- | --- | --- | --- | --- |
| Würzburg | Training | 44 (12) | STN | 60.4 ± 8.2 | 49.84 ± 12.35 | 49.3 ± 24.8 | OFF med / ON stim | 12.6 ± 4.5 | Horn 2017<sup>19</sup> |
| Boston | Test | 100 (30) | STN | 68.4 ± 8.9 | 30.15 ± 13.63 | 27.8 ± 47.5 | ON med / ON stim | 8.5 ± 4.3 | - |
| Boston | Feasibility | 6 (2) | STN<br>GPi | 61.5 ± 5.9 | - | - | OFF med / ON stim | 5.3 ± 4.8 | - |

### The optimization algorithm improves overlap with connectomic targets

In the training cohort, we developed and tuned the optimization algorithm (**Figure 1**) to maximize stimulation volume overlap with symptom-specific connectomic targets, including the above gait circuits (Supplementary Materials 1.1). To ensure our algorithm was performing appropriately in the test cohort, we evaluated if the algorithm increased overlap with the gait network (**Figure 2A**) and fibers (**Figure 2B**), with additional validation on the other symptom targets (Supplementary Materials 1.2). Compared to the patients’ clinical settings, the ‘gait-optimized’ stimulation volumes increased overlap with the gait network (t = 15.8, p < 0.0001) (**Figure 2C, D**), and fibers (t = 17.0, p < 0.0001) (**Figure 2E, F**). The suggested “gait-optimized” settings were more ventral than clinical stimulation volumes for both the gait network (t = 2.07, p = 0.038) (**Figure 2G**) and fibers (t = 2.15, p = 0.041) (**Figure 2H**), although the distribution was often bimodal with activation of both dorsal and ventral contacts (Supplementary Materials 1.3). The ‘gait-optimized’ settings utilized different contacts in 78% of the patients and suggested the use of lower amplitudes than the patients’ clinical settings in 86% of patients (t = 3.91, p < 0.0001) (Supplementary Materials 1.4).

**Figure 1.**
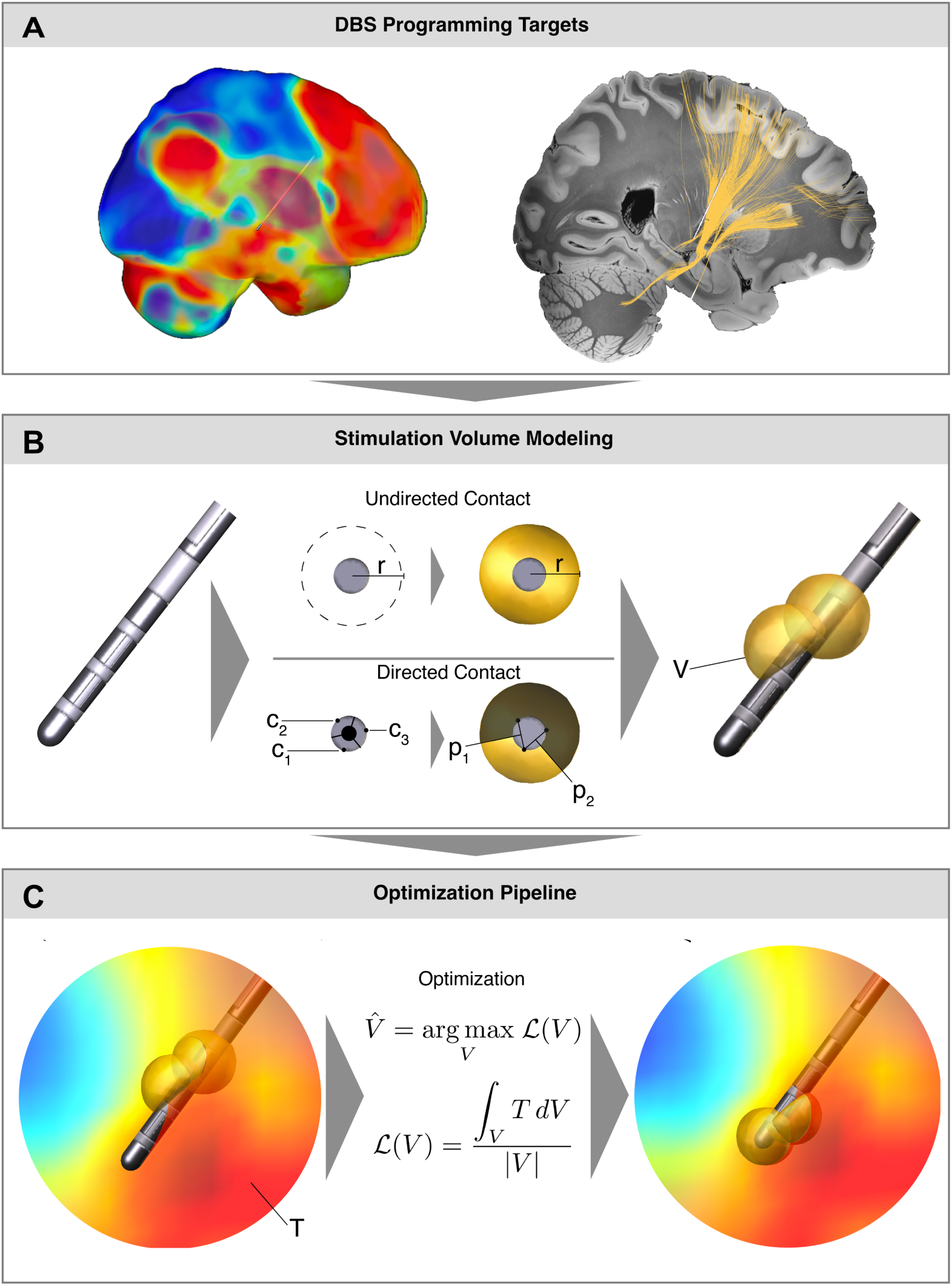
Optimization algorithm to maximize overlap of stimulation volumes with connectomic targets. A) An example of a patient with a DBS electrode in the STN. The gait network (left) and gait fibers (right) are overlaid. (B) The geometric stimulation volume model. The stimulation volume around an undirected contact is approximated as a sphere. The current solves for the radius of the sphere (*r*).^40^ The stimulation volume around a directional contact is approximated as the component of a sphere anterior to two shielding planes (*p*_1_, *p*_2_) between the contact of origin (*c*_1_) and neighboring contacts (*c*_2_, *c*_3_). This solves the pitch, yaw, and roll of the directional stimulation volume. The overall stimulation volume (V) is the union of each contact’s stimulation volumes. C) The stimulation volume from a patient’s clinical DBS settings is shown (left), overlaid on the gait network (T). The optimization function maximizes the overlap of the stimulation volume with the target, yielding the ‘gait-optimized’ stimulation volume (right).

**Figure 2.**
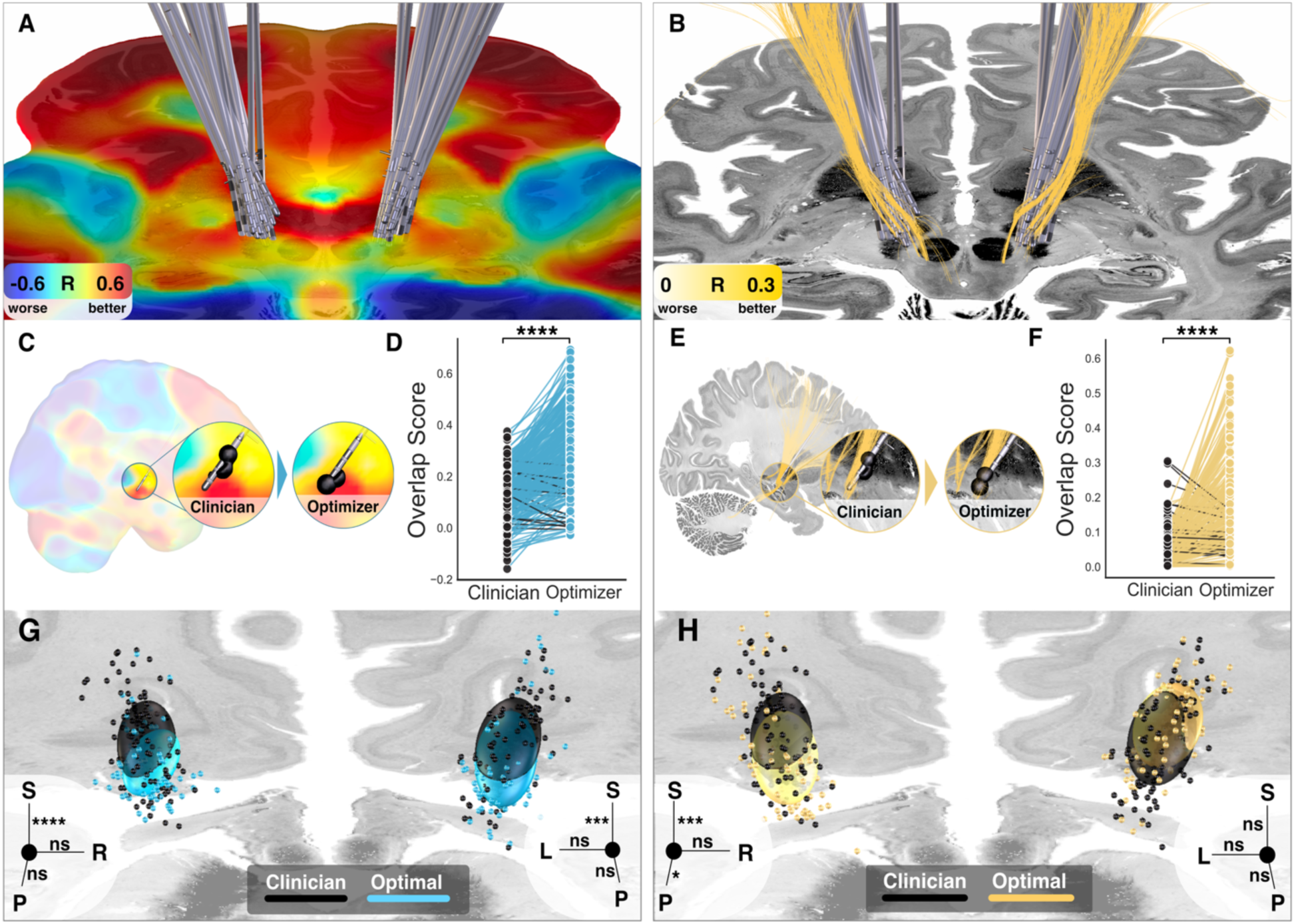
Gait-optimized stimulation volumes increase overlap with gait network and fibers. Test cohort electrode reconstructions are shown overlaid on A) the gait network, and B) the gait fibers. C) An example patient with the clinician’s stimulation volume and corresponding ‘gait-optimized’ stimulation volume from the gait network. D) ‘Gait-optimized’ stimulation volumes significantly increase gait network overlap (t = 27.3, p < 0.0001). E) The same example patient with the clinician’s stimulation volume and corresponding ‘gait-optimized’ stimulation volume from the gait fibers. F) ‘Gait-optimized’ stimulation volumes significantly increase gait fiber overlap (t = 17.0, p < 0.0001). G) The center coordinates of the stimulation volumes for the clinical (black) and ‘gait-optimized’ network settings (blue), with the ellipsoids representing one standard deviation of their axes. The clinical and ‘gait-optimized’ centroids coordinates significantly differed (t = 2.07, p = 0.038), primarily driven by vertical displacement (see inset). H) The stimulation volume center coordinates of the clinical (black) and ‘gait-optimized’ fiber settings (yellow) also significantly differed (t = 2.15, p = 0.041), which was also driven by vertical displacement (see inset). Individual deviations in the x, y, or z coordinates are shown in the axis insets. All p-values are FWE-corrected.

We next investigated if ‘optimized’ stimulation volumes for other symptoms that may more frequently be used to guide clinical DBS programming would better align with the clinical stimulation volumes. Compared to the ‘gait-optimized’ stimulation volumes, the tremor-, rigidity-, bradykinesia-, and UPDRS-III-optimized stimulation volumes were more similar to the clinical stimulation volumes (p < 0.0001) and less distant from the clinical stimulation sites (p < 0.05) (Supplementary Materials 1.5). The other ‘symptom-optimized’ settings selected different contacts from the clinical settings in 48% of patients on average.

### Similarity between ‘gait-optimized’ and clinical stimulation volumes correlates with gait outcomes

In our test cohort, we investigated if similarity between a patient’s actual and ‘gait-optimized’ stimulation volumes was associated with UPDRS-III gait subscore changes one year after DBS for the gait network (**Figure 3A, left**) and fibers (**Figure 3B, left**). Increasing similarity with the ‘gait-optimized’ stimulation volumes was associated with gait improvement one year after DBS for the gait network (r = 0.39, p < 0.0001) (**Figure 3A, right**) and fibers (r = 0.61, p < 0.0001) (**Figure 3B, right**).

**Figure 3.**
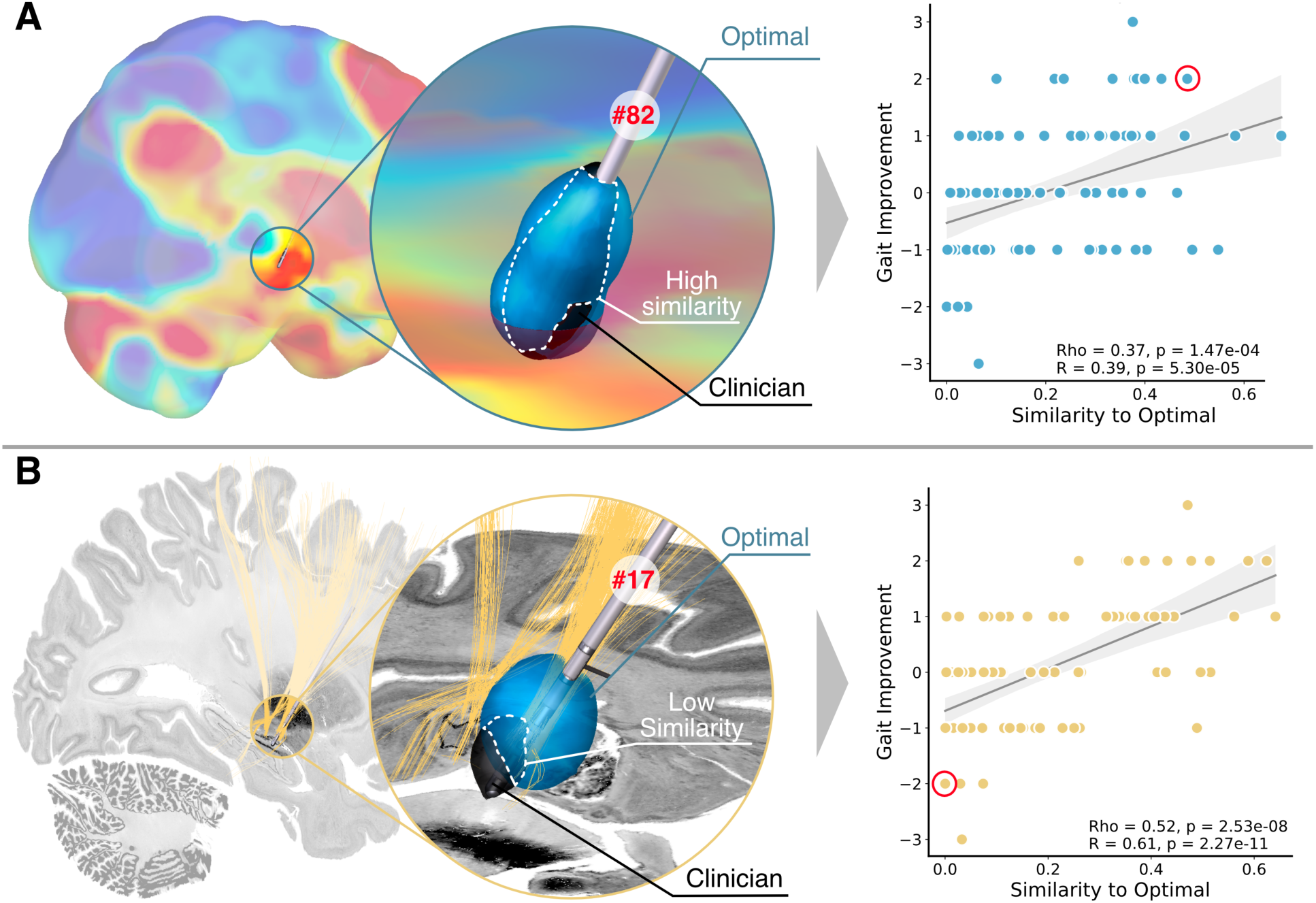
Similarity of clinical stimulation volumes and ‘gait-optimized’ stimulation volumes is associated with gait outcomes. A) A patient with gait improvement and clinical stimulation volumes that were highly similar to the ‘gait-maximal’ stimulation volumes derived from the gait network. For the gait network, similarity of clinical and ‘gait-optimized’ stimulation volumes correlated with gait outcomes after DBS (Rho = 0.37, p < 0.0001). B) A separate patient with gait decline and clinical stimulation volumes that were dissimilar to their ‘gait-optimized’ stimulation volumes derived from the gait fibers. For the gait fibers, similarity of clinical and ‘gait-optimized’ stimulation volumes also correlated with gait outcomes after DBS (Rho = 0.52, p < 0.0001).

Compared to patients with gait improvement (n = 32) after DBS, patients with gait worsening (n = 38) had significantly lower similarity of their clinical stimulation volumes to their ‘gait-optimized’ stimulation volumes (t = 5.25, p = 0.0003) (Supplementary Materials 2.1). These results were gait-specific, as similarity to the ‘gait-optimized’ stimulation volumes was not associated with changes in other symptoms (Supplementary Materials 2.2). The ‘gait-optimized’ settings for the network and the fibers were different (Supplementary Materials 2.3), and the fibers were more strongly associated with gait outcomes (Supplementary Materials 2.4). However, a linear model (R^2^ = 0.31, p < 0.0001) revealed an interaction effect where patients with the highest gait improvements had a high overlap with both gait targets (t = 4.08, p = 0.0001) (Supplementary Materials 2.5).

We next investigated the variance explained by similarity to ‘symptom-optimized’ stimulation volumes for the symptoms more frequently used to guide clinical DBS programming. On average for both networks and fibers, similarity to ‘symptom-optimized’ stimulation volumes derived from both networks and fibers related weakly to outcomes in the UPDRS-III (r = 0.23, p = 0.046) and tremor (r = 0.33, p = 0.021), but not rigidity (r = 0.11, p = 0.62) nor bradykinesia (r = 0.07, p = 0.52) (Supplementary Materials 2.6). On average, similarity of ‘gait-optimized’ stimulation volumes explained more variance in gait than other ‘symptom-optimized’ stimulation volumes could explain variance in their respective symptoms (Δρ^2^ = 0.22, p < 0.0001) (Supplementary Materials 2.7).

### Reprogramming patients to connectomic gait targets

To illustrate how the above information might be used clinically, we reprogrammed 6 Parkinson disease patients (Supplementary Materials 3.1) with a chief complaint of gait dysfunction after STN (n = 3) (**Figure 4A**) or GPi (n = 3) DBS (**Figure 4B**). On ‘gait-optimized’ settings, all 6 patients reported subjective gait improvement (**Figure 4C**). To assess what objective aspect of gait might be driving this subjective change, we compared gait examinations in patients randomly assigned to best-clinical versus ‘gait-optimized’ settings. The ‘gait-optimized’ settings were primarily associated with decreased time spent frozen (78% decrease) and increased walk speed (20% increase) (**Figure 4D**). However, this cohort is too small for meaningful statistical inference. On ‘gait-optimized’ settings, tremor recurred in 5 patients, and 1 had stimulation-induced dyskinesias. No other adverse effects were identified, including paresthesia, plegia, pain, vertigo, and ataxia. Compared to clinical settings, the ‘gait-optimized’ settings activated different electrode contacts in 83% of patients, increased overlap with the gait targets, but reduced overlap with the UPDRS-III, tremor, rigidity, and bradykinesia targets (Supplementary Materials 3.2).

**Figure 4.**
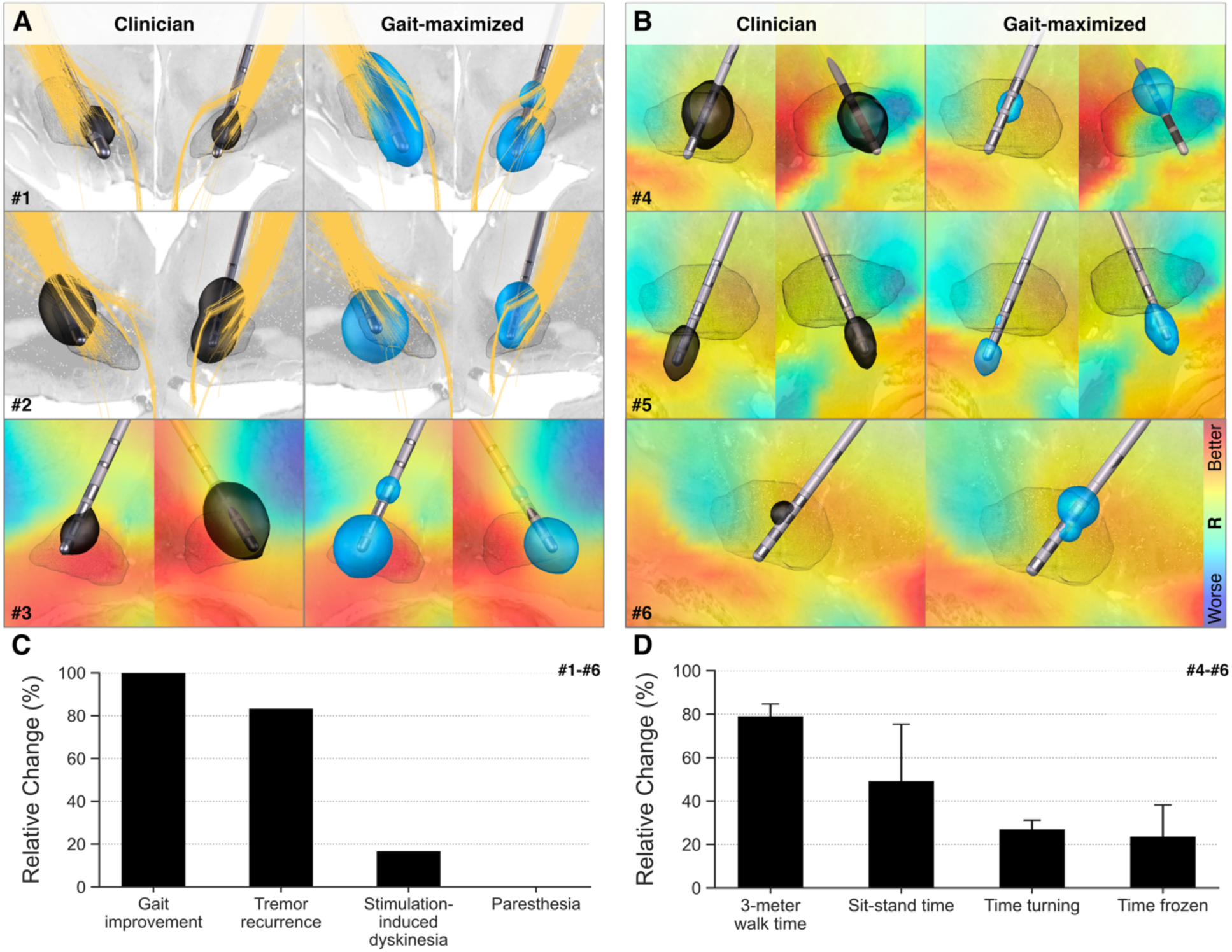
Prospective reprogramming of patients with gait dysfunction to ‘gait-optimized’ DBS parameters. A) The three patients with STN DBS who were reprogrammed from best clinical (black) to ‘gait-optimized’ (blue) settings. Patients 1 and 2 were programmed to the gait fibers, while patient 3 was programmed to the gait network. B) The three patients with GPi DBS who were reprogrammed from best clinical (black) to ‘gait-optimized’ (blue) settings. The gait fibers were not present in the GPi, and all patients were programmed to the gait network. C) On ‘gait-optimized’ settings, 100% of patients reported gait improvement although 82.5% reported tremor recurrence on the new gait settings. One patient (12.5%) had transient stimulation-induced dyskinesias, and no patients reported paresthesia nor other adverse effects on ‘gait-optimized’ settings. D) For the four single-blinded patients who randomly received clinical or ‘gait-optimized’ settings, the ‘gait-optimized’ settings were associated with 20% increase walk speed (t = 3.69, p = 0.066), 53% less time spent in the sit-to-stand transition (t = 1.94, p = 0.19), 86% less time spent turning (t = -17.5, p = 0.0032), and 78% less time frozen (t = 17, p = 0.0033).

## Discussion

We developed a neuroimaging-based algorithm which uses the location of a patient’s implanted electrode to suggest DBS settings that maximizes overlap of DBS stimulation volumes with brain networks and/or fiber tracts. This algorithm increased stimulation volume overlap with two recent connectomic targets for gait dysfunction,^15,16^ resulting in stimulation volumes that differed significantly from clinical settings. Retrospectively, increased similarity of a patient’s actual DBS stimulation volume to the ‘gait-optimized’ stimulation volumes was associated with improved gait while dissimilarity was associated with gait worsening. In the handful of prospective patients, ‘gait-optimized’ settings were associated with gait improvement.

### Optimization algorithm for use with connectomic targets

While prior studies have used an algorithm to optimize DBS parameters to intersect white matter fiber tracts^13,16^ we are aware of only one study that has tried to intersect a functionally connected brain network, and this study used expert visual inspection rather than a fitting algorithm.^18^ Our algorithm, Stim PyPer, can identify optimal DBS settings on the location of an individual patients implanted electrode designed to intersect either a set of fiber tracts or a functional brain network, increasing versatility for different types of brain circuit targets^15,16,19,20^ or potentially local sweet-spots.^21,22^

We did find that fiber-based optimization may be more strongly related to clinical outcomes than network-based stimulation. However, there was an interaction such that patients with the best gait outcomes had high overlap with both circuit types, and both targets were associated with gait improvement in the feasibility cohort. Taken together, these results suggest network- and fiber-based targets capture partially independent aspects of gait-related circuitry, which may provide complementary information for DBS programming.

Our results suggest that some symptoms may benefit more from image guidance algorithms than others. For example, the optimal settings for tremor or bradykinesia were similar to the clinically selected settings. For these symptoms, the main value of image guidance may be in saving the clinician time.^23^ Image guided programming may have a larger role in symptoms which are harder to program to in a clinical setting, due to delayed effects or difficulty in measuring symptom changes. Examples within Parkinson disease may include gait,^2^ depression,^24^ or cognitive decline.^25^

### Gait-optimized DBS settings and gait outcomes

Gait dysfunction is a particularly challenging symptom to treat in Parkinson disease,^1,2,26^ and there is concern that DBS can both impair or benefit gait depending on the DBS settings.^1,2,27,28^ Our results add support to these observations. We find the ‘gait-optimized’ stimulation volumes differ significantly from clinical stimulation volumes. Retrospectively, patients with clinical stimulation volumes that were serendipitously similar to their ‘gait-optimized’ stimulation volumes tended to have gait improvement. However, patients with low similarity to their ‘gait-optimized’ stimulation volumes tended to have gait worsening after DBS. In the handful of prospectively reprogrammed patients, gait and freezing improved on their ‘gait-optimized’ settings. These results support the findings that DBS programming may both improve or impair gait,^1^ which may be partially driven by how much DBS stimulation volumes overlaps circuitry associated with gait dysfunction. ^2,27^ We find the overall centroid of stimulation volumes for gait are more ventral than clinical settings, however a simple heuristic that moves stimulation more ventrally may not be adequate for gait improvement. For example the gait-optimized settings often had both ventral and dorsal stimulation volumes despite an overall ventral shift. This is consistent with prior sweet-spot studies which found dorsal stimulation may be associated with gait benefit,^12^ but suggests that simultaneous ventral involvement may also be beneficial.

### Symptom-specific circuit engagement and DBS trade-offs

Our results also suggest a symptom-specific relationship between gait and the other symptoms of Parkinson disease commonly treated with DBS. ‘Gait-optimized’ stimulation volumes differed significantly from best clinical settings, and were associated with reduced stimulation volume overlap with circuitry linked to tremor, rigidity, and bradykinesia. ‘Symptom-optimized’ stimulation volumes for other symptoms were more similar to clinical stimulation volumes. Additionally, similarity to ‘gait-optimized’ stimulation volumes explained more gait variance than any other ‘symptom-optimized’ stimulation volume explained of their respective symptoms (i.e. similarity to ‘gait-optimized’ stimulation volumes explained more gait variance than ‘rigidity-optimized’ stimulation volumes explained rigidity variance). Taken together, these findings suggest the symptoms commonly treated with DBS are already near-optimally programmed on clinical settings, while gait may be under-optimized.

When we tested the feasibility of circuit-guided reprogramming for gait dysfunction in a small cohort, we found that the clinical settings were associated with tremor control but impaired gait, whereas ‘gait-optimized’ settings were associated with improved gait but tremor recurrence. These findings suggest that ‘optimization’ of DBS to specific symptoms may result in recurrence of others. While reprogramming for gait dysfunction may be feasible, future study design will need to incorporate the possibility that both tolerability and blinding may be affected as symptoms like tremor may recur.

### Limitations

Our optimization model is subject to limitations. To achieve optimization within clinically reasonable timeframes, we simplify biophysical modelling by using geometric approximations for stimulation volume models. Alternative stimulation volume models may more appropriately represent the complex biophysical effects of DBS.^29–32^ Additionally, our optimization algorithm may be limited when applied to DBS electrodes with pulse generators that cannot specify unique currents across each contact. Further, while clinicians can avoid adverse effects in real time, our optimization algorithm is unaware of what regions might cause adverse effects unless these regions are specifically incorporated.

The retrospective component of this study is also subject to limitations. As with all retrospective studies, causality must be interpreted with caution due to lack of ability to control for confounds. Additionally, while the discovery cohort was evaluated in the off-med state, the testing cohort was evaluated in the on-med state. While this makes interpretation more challenging, it suggests the effects of DBS upon gait may be of sufficient effect size to be detected despite being on medications. Lastly, while we did investigate the effect of ‘symptom-optimized’ stimulation upon other symptoms, we found these relationships were small, which may be due to the on-medication state.

Further, the small feasibility study of circuit-based gait reprogramming limited in its sample size and open label design. Many other studies have shown DBS can influence gait under similar circumstances,^6,33–35^ but have had mixed translation to clinical trials and practice. To know the actual effect size of ‘gait-optimized’ DBS, a proper randomized controlled trial comparing ‘gait-optimized’ to clinical programming is required.

## Methods

### Ethics Statement

This study was conducted in accordance with ethical standards and approved by the Institutional Review Board (protocol #2021P002226) of the Brigham and Women’s Hospital and Harvard Medical School, Boston, Massachusetts, as well as the Institutional Review Board (protocol #2015P00028) of Beth Israel Deaconess Medical Center, Boston, Massachusetts.

### Subjects

The study included 3 cohorts of patients with Parkinson disease who underwent DBS in Würzburg, Germany, or Boston, USA. Cohort characteristics are summarized in Table 1. The training cohort (n = 44) from Würzburg^18,19^ used Medtronic 3387 and 3389 electrodes with average parameters of 150 Hz, pulse width 50 μs, and 3.5 mA. The test cohort (n = 100) from Boston used Medtronic 3387, 3389, B33005, B33015; Boston Scientific DB-2202; and Abbott 6172 electrodes with average parameters of 130 Hz, 60 μs, and a range of current amplitudes. The feasibility cohort (n = 6) used Medtronic 3387 electrodes with average parameters of 130 Hz, pulse width 60 μs, and a range of current amplitudes. All gait changes were pre-operative versus one-year postoperative changes. Informed consent was achieved for all subjects.

### Connectomic symptom-specific targets

We chose two established and validated normative gait-specific targets, a gait network derived with resting state fMRI and fibers derived from structural connectivity^16^ (**Figure 1A**).^15^ Additional tract-specific targets were available for specific symptoms of Parkinson including tremor, rigidity, bradykinesia, and axial symptoms,^16^ as well as one prior tremor brain network.^20^ Using the training cohort, symptom networks were generated for the UPDRS-III, rigidity, and bradykinesia which correspond directly to the tract-based targets.^16^ Network mapping consisted of deriving the whole-brain functional connectivity profile of the DBS sites,^36^ and correlating these connections with outcomes in the tremor, rigidity, bradykinesia, and axial symptom categories to generate the additional symptom-specific networks.

### Optimization software for maximization of stimulation volumes to connectomic targets

We developed a fast geometric optimization framework to identify patient-specific DBS stimulation settings that maximize overlap with symptom-specific connectomic targets. Stimulation volumes were approximated using closed-form geometric models derived from electrode contact coordinates (Supplementary Materials 4.1). Functional network targets and tract-based targets were converted into a common voxelwise space using connectivity inversion and tract-density estimation, respectively (Supplementary Materials 4.2). Optimization maximized the mean target overlap within the stimulation volume across all active contacts while penalizing unsafe current levels (Supplementary Materials 4.3). Hyperparameters were selected by grid search in the training cohort (Supplementary Materials 4.4). Initial amplitudes were estimated using a multistart strategy and triangulation of potential maxima (Supplementary Materials 4.5).

### Evaluation of stimulation volumes generated from optimization algorithm

To investigate if the optimized stimulation volumes increased overlap with symptom-specific targets, we generated the ‘symptom-optimized’ stimulation volumes and measured their overlap with the given symptom-specific target. For a null representation of reasonable stimulation volumes, we repeated the process for the patients’ current clinical stimulation volumes. These overlap values were compared with a paired t-test.

Additional metrics, such as their central coordinates and Dice coefficients were also compared.

### Relationship of ‘gait-optimized’ stimulation volumes with gait symptoms

To assess if the ‘gait-optimized’ stimulation volumes might be associated with gait outcomes, we measured the similarity (Dice coefficient) between a patient’s clinical stimulation volumes and their ‘gait-optimized’ stimulation volumes. We correlated this similarity value to gait change one year after surgery, as measured by the gait subscale of the UPDRS-III.

### Feasibility of reprogramming of gait-specific targets in Parkinson Disease DBS

We reprogrammed several patients to optimize their stimulation volumes to the gait-specific network and fibers. Within Boston DBS clinics, patients with chief complaints of gait dysfunction after DBS were identified (Supplementary Materials 3.1). An expert movement disorders neurologist reprogrammed DBS settings to ‘gait-optimized’ parameters. Outcomes were subjectively evaluated by the patients and objectively evaluated by neurological examination. Prospective gait evaluations of patients on ‘gait-optimized’ DBS settings were normalized to their gait evaluations on best clinical settings. Lower normalized scores represented less time required to walk or less time frozen, representing improvement.

### Statistics statement

Statistical analyses were conducted in Python 3.10 using Statsmodels 0.14.0, Nilearn 0.10.1, and Scikit-learn 1.3.0.^37–39^ Descriptive statistics are presented as mean ± standard deviation. Correlations used Spearman correlations as appropriate for ordinal data. Gait outcomes in prospective patients were compared to the null hypothesis of baseline gait performance using one sample t-tests. Superiority comparisons were done by bootstrapped estimation of correlation values. The resulting distributions were compared with a paired t-test and 95% confidence intervals.

## Data Availability

All data produced in the present study are available upon reasonable request to the authors

## Acknowledgements

M.M.R. is supported by the German Research Foundation (DFG; – Project-ID 424778381) and the German Federal Ministry of Education and Research (BMBF; Project-ID 01KG2032_DIPS). A.H. was supported by the Schilling Foundation, the German Research Foundation (Deutsche Forschungsgemeinschaft, CRC-1451, 431549029 and CRC-1270 ELAINE, 3–299150580).

