## Supplementary Materials for "Towards connectome-guided optimization of deep brain stimulation for gait dysfunction"

**Connectome-guided optimization of deep brain stimulation for gait dysfunction differs from clinical settings and correlates with gait outcomes**

SUPPLEMENTARY MATERIALS

**Table of Contents**

- S1: Ability of optimized settings target symptom-specific circuitry
  - S1.1: Tuning in the training cohort
  - S1.2: Overlap with symptom-specific targets after optimization to other connectomic symptom targets
  - S1.3: How gait-optimized stimulation volumes differ from clinical stimulation volumes
  - S1.4: Current allocated by algorithm differs from clinical current allocation
  - Stimulation sites from optimization to other connectomic symptom targets
- S2: Relating optimized settings to clinical outcomes
  - S2.1: Clinical stimulation volume similarity to ‘gait-optimized’ stimulation volumes relates to gait outcomes
  - S2.2: Similarity to ‘gait-optimized’ stimulation volumes relates specifically to gait outcomes
  - S2.3: Optimal stimulation parameters for networks and fibers diverge
  - S2.4: Optimal gait outcomes are a joint function of increased similarity to both network- and fiber-based ‘gait-optimized’ stimulation volumes
  - S2.5: Optimal gait outcomes are a joint function of increased similarity to both network- and fiber-based ‘gait-optimized’ stimulation volumes
  - S2.6: Relationship between outcomes and optimal stimulation programs for other PD symptom domains
  - S2.7: Similarity to ‘symptom-optimized’ stimulation volumes explains variance differentially
- S3: Prospective patients reprogrammed to ‘gait-optimized’ settings
  - S3.1: Detailed clinical features of the prospective cohort
  - S3.2: Prospective cohort ‘gait-optimized’ DBS settings
- S4: Optimization algorithm methodology
  - S4.1: Fast geometric approximation of stimulation volumes to enable iteration
  - S4.2: Converting functional- and tract-based symptom-specific targets into optimization targets
  - S4.3: Optimization of stimulation volumes to symptom-specific connectomic targets
  - S4.4: Hyperparameter tuning
  - S4.5: The initial guess for stimulation volume optimization

**Supplementary Materials 1: Algorithm Development**

S1.1. Tuning in the training cohort

To reach goals of the present study, development of an optimization framework was needed which would suggest DBS parameters that lead to stimulation volumes that maximally engage a given map. The aim of the optimizer was defined in such a way, that it would maximize overlap with either binary or nonbinary target maps (Supplementary Methods). To evaluate reliable convergence of the optimizer to global maxima (i.e. convexity), we first validated the algorithm using a synthetic target, where it consistently converged on the global optimum (p < 0.0001). When electrodes were placed within negative regions of this synthetic environment, the algorithm shut them off (p < 0.0001). However, the algorithm may fail to find global maxima as the target becomes more complex (i.e. non-convexity), such as may occur with real connectomic targets.

To evaluate convexity when using real connectomic targets, we evaluated the optimizer’s results using the gait-specific brain network and fibers. In this environment, the algorithm originally generated highly variable results across repeated runs even within the same patient (Supplementary Figure 1A). To address this non-convexity, we implemented a first guess strategy based on triangulating the local maxima around each contact, along with a multi-start approach (Supplementary Figure 1B). This approach reduced variance across runs and improved the overlap between the algorithm-suggested stimulation volume and the neuromodulation target (p = 0.0008, Supplementary Figure 1C). We next performed a grid search to tune the algorithm’s hyperparameters, which maximized overlap of the stimulation volume with both functional- and tract-based targets (Supplementary Figure 1D).

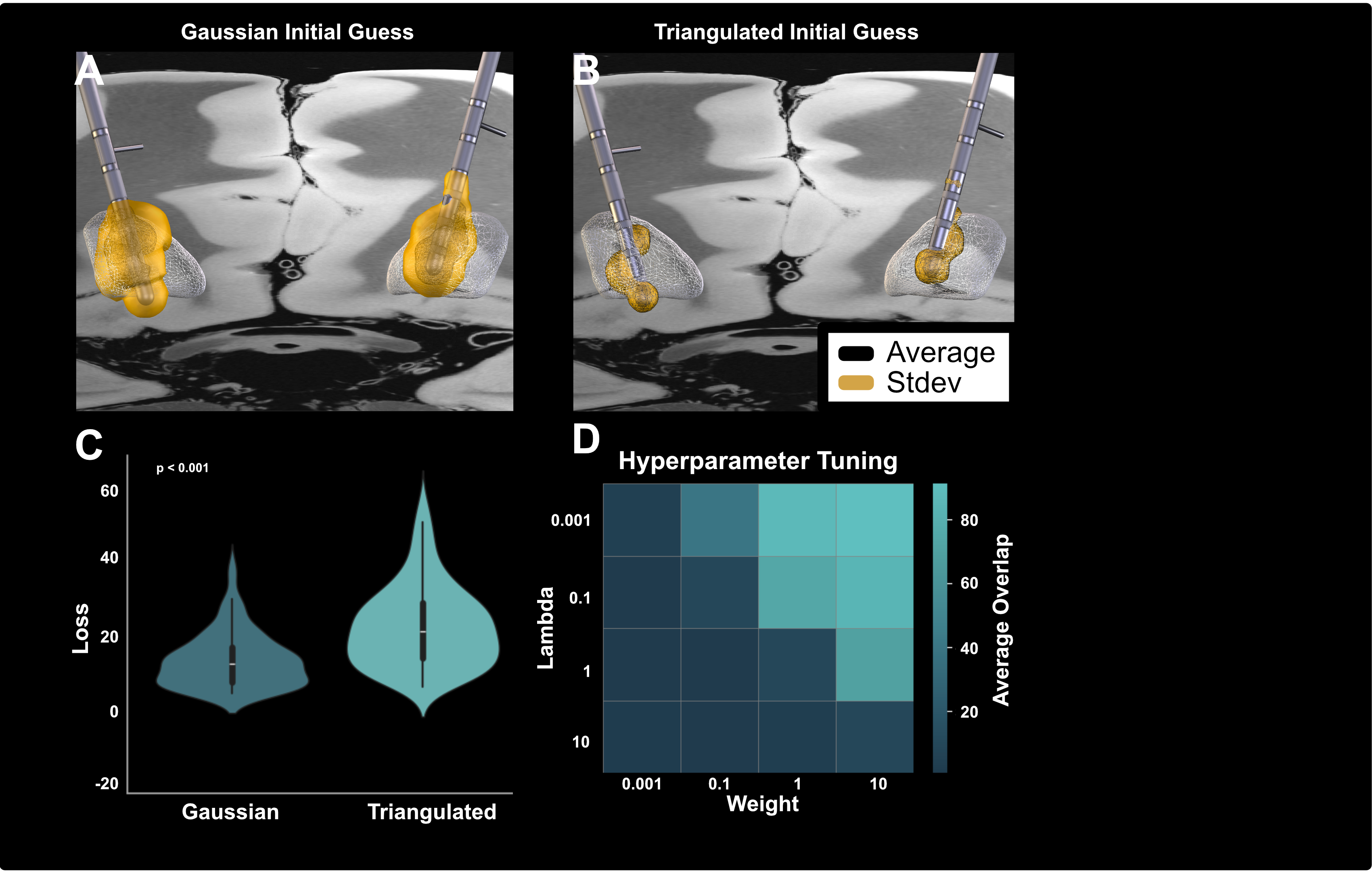

**Supplementary Figure 1. Comparison of initialization strategies and hyperparameter tuning for the DBS optimization algorithm.** A) Gaussian initial guess and B) triangulated initial guess visualized on patient MRI, showing the average (black inner mesh) and standard deviation (gold outer mesh) of the resulting stimulation volumes across resamples. The black inner mesh and gold outer mesh overlap in (B), representing a mean with a very narrow standard deviation compared to (A). C) Distribution of loss values across patients for Gaussian and triangulated initializations, with Gaussian yielding significantly lower loss (p < 0.001). D) Heatmap of overlap scores (average between optimizing to fMRI- and tract-derived targets) across combinations of hyperparameters (weight and lambda), demonstrating regions of optimal performance.

We next generated symptom-specific networks (gait, tremor, rigidity, and bradykinesia) from the training cohort and evaluated if this algorithm could improve stimulation-volume overlap with each of them. Compared to expert clinical settings from the training cohort, the tuned algorithm successfully increased overlap of the stimulation volume with the network (all p < 0.0001) (Supplementary Figure 2), and fiber targets (all p < 0.0014) (Supplementary Figure 3). This circular analysis was completed within the training dataset for evaluation of performance prior to evaluation in the test dataset.

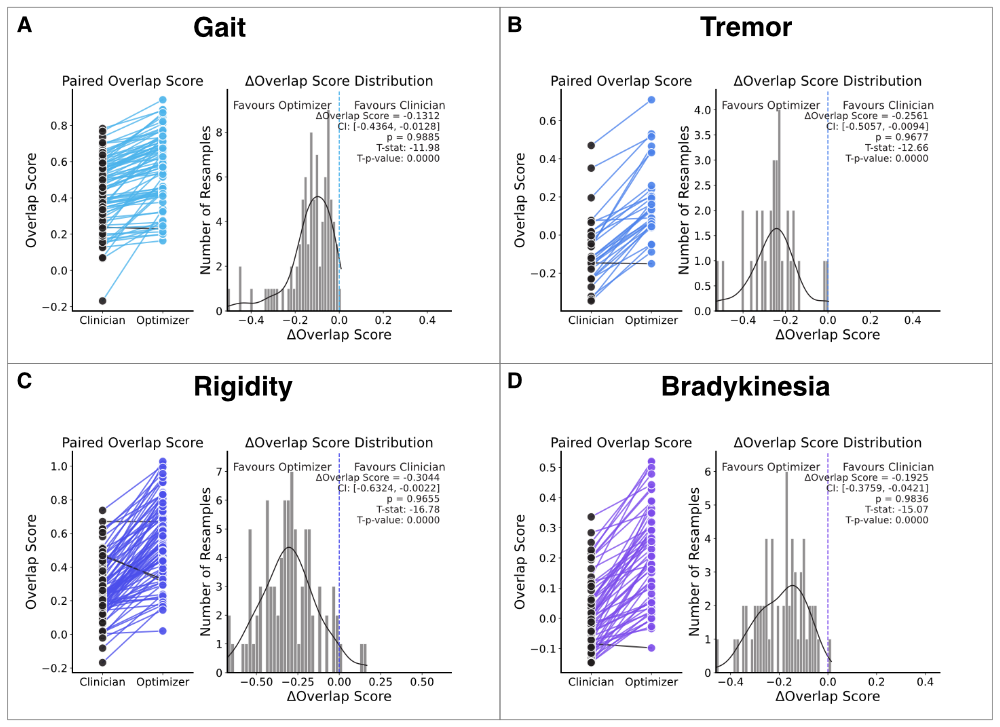

**Supplementary Figure 2. The optimization algorithm successfully reprograms DBS electrodes to target fMRI-derived motor symptom-specific targets in the training cohort.** A) Optimized stimulation volumes significantly improve overlap with the gait target compared to clinician-selected volumes (t = -11.98, p < 0.0001). B) Optimized stimulation volumes significantly improve overlap with the tremor target (t = -12.66, p < 0.0001). C) Optimized stimulation volumes significantly improve overlap with the rigidity target (t = -16.78, p < 0.0001). D) Optimized stimulation volumes significantly improve overlap with the bradykinesia target (t = -15.07, p < 0.0001). Left panels show paired overlap scores between clinician- and optimizer-selected programs for individual patients. Right panels show distributions of overlap score differences (ΔOverlap) from resampling analyses, with confidence intervals and p-values indicating statistical significance in favor of the optimizer.

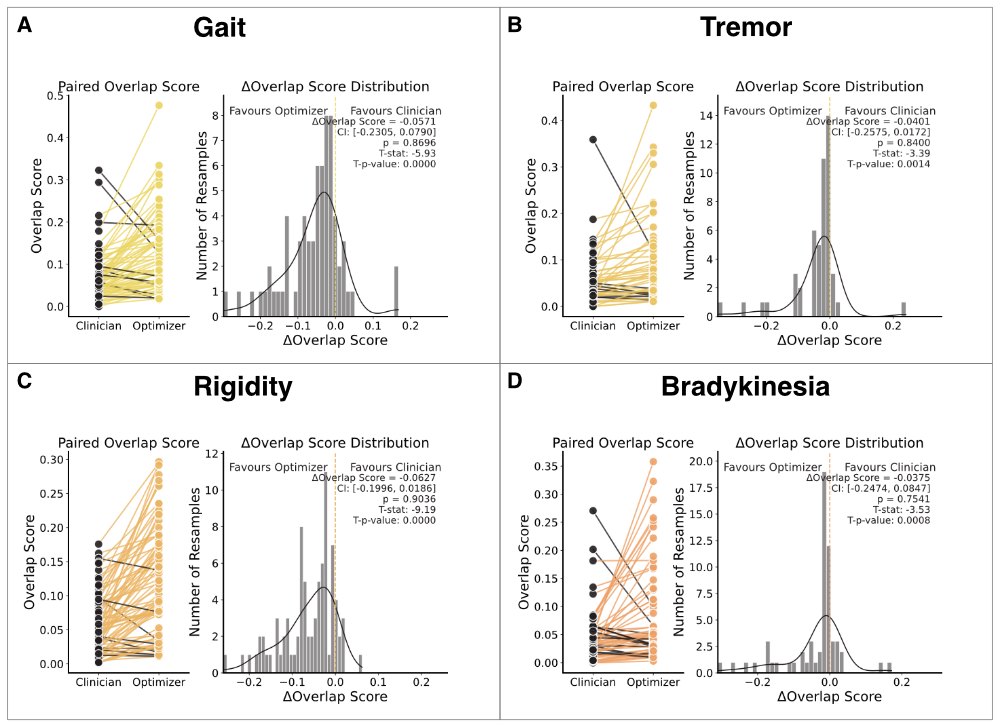

**Supplementary Figure 3. The optimization algorithm successfully reprograms DBS electrodes to target tract-derived motor symptom-specific targets in the training cohort.** A) Gait: Optimized stimulation volumes significantly improve overlap with the gait target compared to clinician-selected volumes (t = -5.93, p < 0.0001). B) Tremor: Optimized stimulation volumes significantly improve overlap with the tremor target (t = -3.39, p = 0.0014). C) Rigidity: Optimized stimulation volumes significantly improve overlap with the rigidity target (t = -9.19, p < 0.0001). D) Bradykinesia: Optimized stimulation volumes significantly improve overlap with the bradykinesia target (t = -3.53, p = 0.0008). Left panels show paired overlap scores between clinician- and optimizer-selected programs for individual patients. Right panels show distributions of overlap score differences (ΔOverlap) from resampling analyses, with confidence intervals and p-values indicating statistical significance in favor of the optimizer.

S1.2. Overlap with symptom-specific targets after optimization to other connectomic symptom targets
We next wanted to investigate if the optimization algorithm’s ability to increase overlap with the symptom-specific target was specific to gait, or could generalize to other symptoms. In our test cohort, which was not used to tune the parameters nor build the targets, we generated ‘symptom-optimized’ stimulation volumes for the overall UPDRS-III, tremor, rigidity, and bradykinesia. This was done with both networks and fibers. The ‘symptom-optimized’ stimulation volumes significantly increased overlap with the symptom-networks in the test cohort (largest p < 0.0001) (Supplementary Figure 4), as well as the symptom-fibers in the test cohort (largest p < 0.0001) (Supplementary Figure 5).

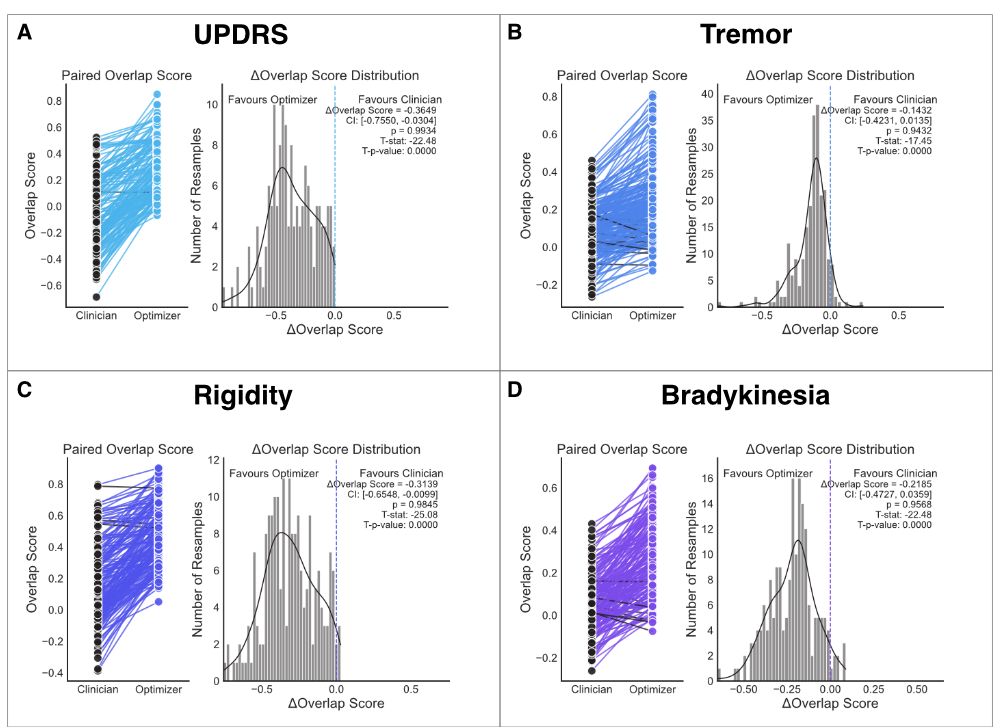

**Supplementary Figure 4. The optimization algorithm successfully reprograms DBS electrodes to target fMRI-derived motor symptom-specific targets in the test cohort.** A) UPDRS: Optimized stimulation volumes significantly improve overlap with the UPDRS target compared to clinician-selected volumes (t = -22.48, p < 0.0001). B) Optimized stimulation volumes significantly improve overlap with the tremor target (t = -17.45, p < 0.0001). C) Optimized stimulation volumes significantly improve overlap with the rigidity target (t = -25.08, p < 0.0001). D) Optimized stimulation volumes significantly improve overlap with the bradykinesia target (t = -22.48, p < 0.0001). Left panels show paired overlap scores between clinician- and optimizer-selected programs for individual patients. Right panels show distributions of overlap score differences (ΔOverlap) from resampling analyses, with confidence intervals and p-values indicating statistical significance in favor of the optimizer

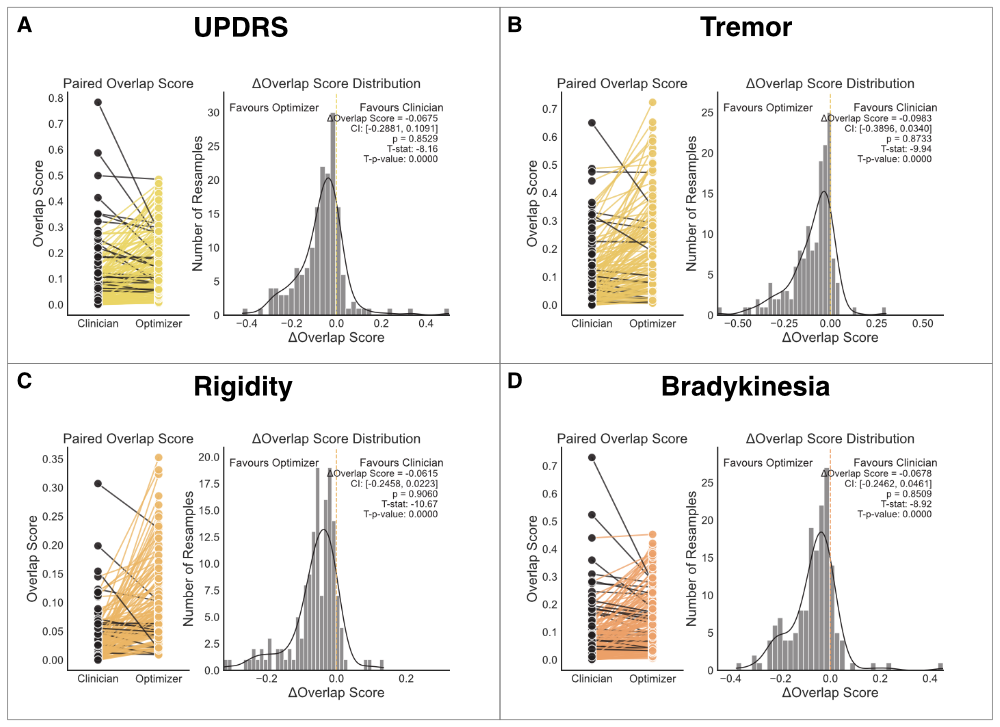

**Supplementary Figure 5. The optimization algorithm successfully reprograms DBS electrodes to target tract-derived motor symptom-specific targets in the test cohort.** A) ‘UPDRS-optimized’ stimulation volumes significantly improve overlap with the UPDRS fibers compared to clinical stimulation volumes (t = -8.16, p < 0.0001). B) ‘Tremor-optimized’ stimulation volumes significantly improve overlap with the tremor fibers compared to clinical stimulation volumes (t = -9.94, p < 0.0001). C) ‘Rigidity-optimized’ stimulation volumes significantly improve overlap with the rigidity fibers compared to clinical stimulation volumes (t = -10.67, p < 0.0001). D) ‘Bradykinesia-optimized’ stimulation volumes significantly improve overlap with the bradykinesia fibers compared to clinical stimulation volumes (t = -8.92, p < 0.0001). Left sub-panels show paired overlap scores between clinician stimulation volumes and ‘symptom-optimized’ stimulation volumes, paired within patients. Right panels show distributions of overlap score differences (Δ Overlap) from resampling analyses, with confidence intervals and p-values indicating statistical probability of the ‘symptom-optimized’ stimulation volumes having higher overlap with symptom-specific fibers than the clinical stimulation volumes.

S1.3. How gait-optimized stimulation volumes differ from clinical stimulation volumes

We investigated how different ‘gait-optimized’ settings for the network and tract targets were to the current clinical settings. Compared to clinical settings, the optimizer drove stimulations more inferior, which was significant when considering the hemispheres independently or together (Supplementary Figure 6). This was not due to a simple downward displacement, as passing 2 milliamps of current at the lowest contact did not increase overlap with the gait targets compared with clinician settings (t = 1.52 p = 0.51), suggesting a combination of vertical displacement, contact directionality, and current magnitude contributed to the difference in overlap with the gait network and fibers.

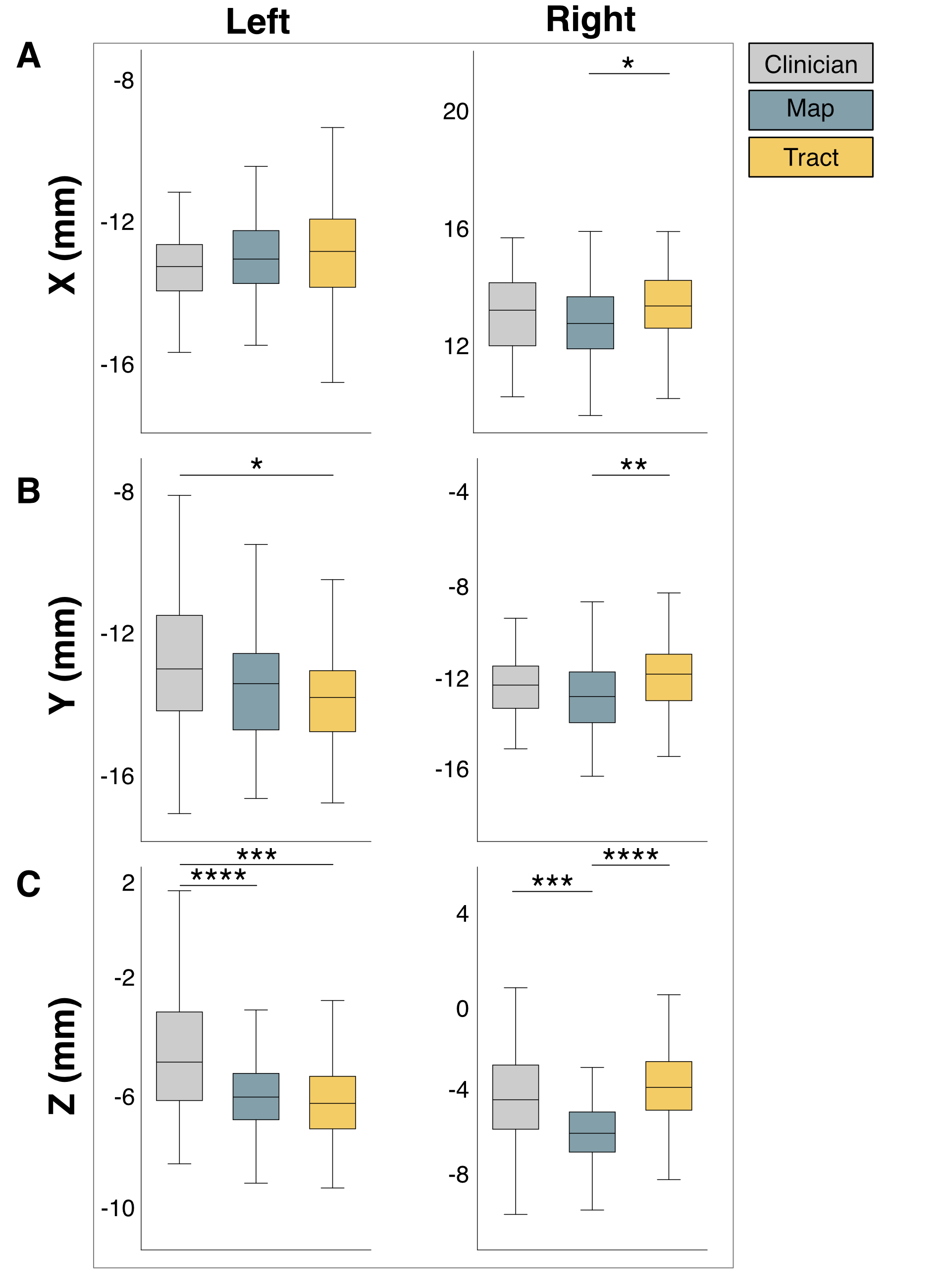

**Supplementary Figure 6. Stimulation volume centroid coordinates across programming methods.** Boxplots of the median and interquartile range of stimulation volume locations in MNI space (mm) along the X (medial-lateral), Y (anterior-posterior), and Z (superior-inferior) axes for the left (left column) and right (right column) hemispheres. Within each side and axis, the three programs were compared, including the clinician- (gray), network- (blue), and fiber-based settings (yellow). (A) Along the X-axis, no significant differences were found in the left hemisphere, while in the right hemisphere, map-based and tract-based programs diverged (t = 2.73, p_fwe_ = 0.04). (B) Along the Y-axis, a significant difference was observed in the left hemisphere between clinician and fiber-based programs (t = 2.82, p_fwe_ = 0.03), and in the right hemisphere between network- and fiber-based programs (t = 3.13, p_fwe_ = 0.01). (Z) Along the Z-axis, strong effects were observed bilaterally: in the left hemisphere, clinician vs. map (t = 5.04, p_fwe_ < 0.0001) and clinician vs. tract (t = 4.14, p_fwe_ = 0.0003); in the right hemisphere, clinician vs. network (t = 3.32, p_fwe_ = 0.006) and network vs. fibers (t = 5.09, p_fwe_ < 0.0001). Legend: map (network), tract (fibers).

S1.4. Current allocated by algorithm differs from clinical current allocation

Within our test cohort, we compared the allocation of electrical current across each electrode’s contacts by the optimizer and clinicians (Supplementary Figure 7, left). A two-way ANOVA found a significant difference in allocation of current across the individual contacts, comparing the optimizer and clinician (F = 45.16, p < 0.0001). We then evaluated the total current passed across the electrode (sum across contacts), and found the optimizer allocated significantly less current than clinicians (t = 3.91, p = 0.0001) (Supplementary Figure 7, right). Although the clinical settings exceeded 5mA roughly a third of the time, the optimizer never exceeded 5mA of total current.

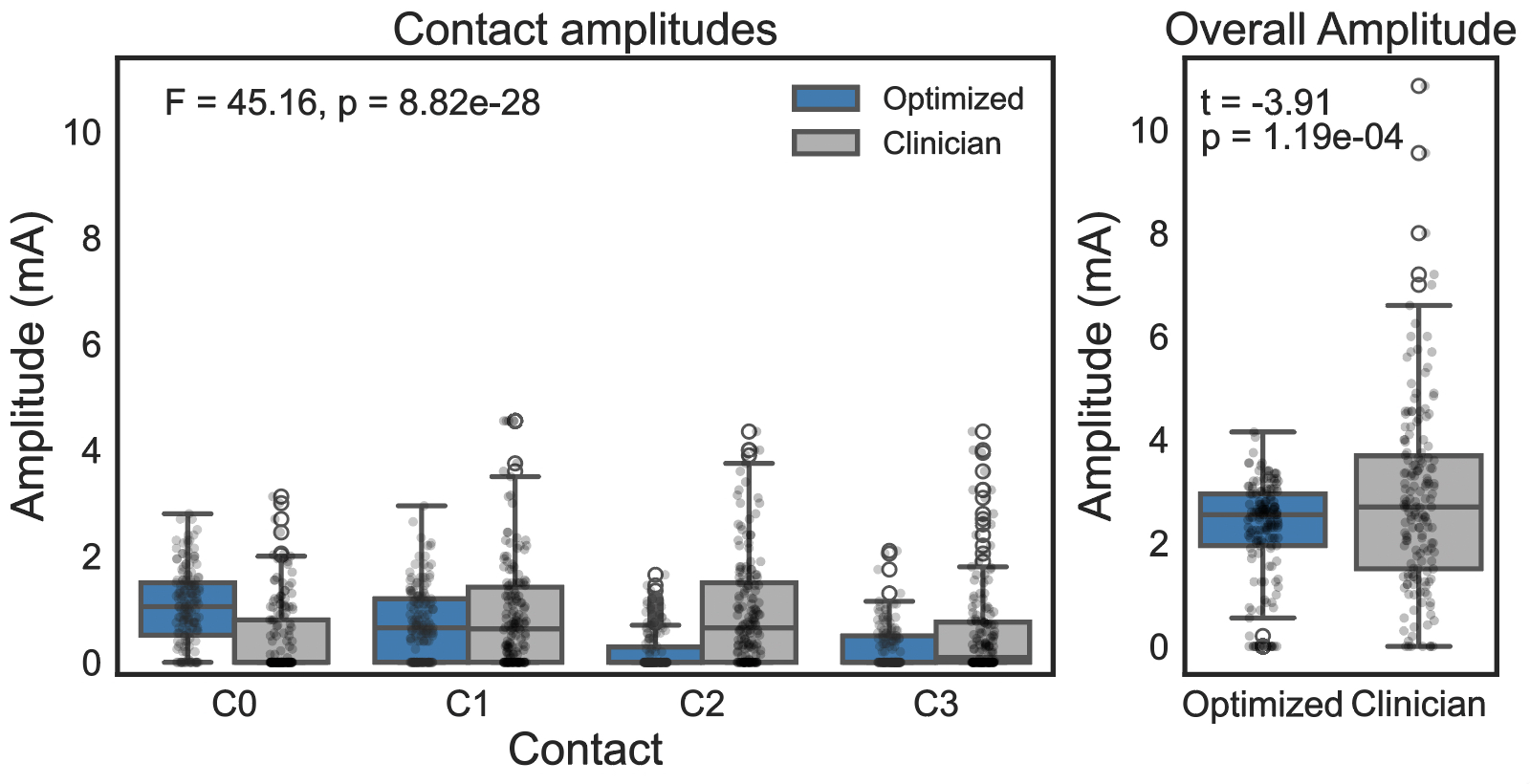

**Supplementary Figure 7. Distribution of stimulation amplitudes assigned by the optimization algorithm compared with clinician-selected settings.** The left panel shows current amplitudes across individual electrode contacts (C0–C3). A two-way analysis of variance comparing allocation patterns between optimizer-derived and clinician-selected programs revealed a significant difference in how current was distributed across contacts (F = 45.16, p < 0.0001). The right panel shows the total stimulation amplitude delivered by each program (sum of currents across contacts). The optimizer assigned significantly lower total current than clinician-selected programs (t = −3.91, p = 0.0001). Each point represents an individual stimulation program; boxplots indicate the median and interquartile range with whiskers extending to 1.5 times the interquartile range. Optimized settings from both the networks and fibers are evaluated relative to clinical settings.

S1.5. Stimulation sites from optimization to other connectomic symptom targets

Within our test cohort, we generated ‘symptom-optimized’ stimulation volumes for the overall UPDRS-III, tremor, rigidity, and bradykinesia. This was done with both networks and fibers. For each stimulation volume, we extracted the average centroid coordinate of the binary stimulation volumes. For each symptom, we then measured the distance of this coordinate to the clinical stimulation site coordinates. We then compared the distance of the ‘gait-optimized’ stimulation volumes from the clinical stimulation volumes to the distance of the ‘symptom-optimized’ stimulation volumes from the clinical stimulation volumes. Overall, the ‘gait-optimized’ stimulation volumes were significantly farther from the clinical stimulation sites than the other ‘symptom-optimized’ stimulation volumes for both networks (t = 2.09, p = 0.038) and fibers (t = 5.89, p < 0.0001). We also compared the Dice coefficient of each patient’s ‘symptom-optimized’ stimulation volume to the corresponding clinical stimulation volume, and found that the ‘gait-optimized’ stimulation volumes had significantly lower overlap with the clinical stimulation volumes for both the networks (t = 7.81, p < 0.0001) and fibers (t = 11.76, p < 0.0001).

We also compared the Dice coefficient of each patient’s ‘symptom-optimized’ stimulation volume to the corresponding clinical stimulation volume, and found that the ‘gait-optimized’ stimulation volumes had significantly lower overlap with the clinical stimulation volumes for both the networks (t = 7.81, p < 0.0001) and fibers (t = 11.76, p < 0.0001).

**Supplementary Materials 2: Association between ‘symptom-optimized’ stimulation and clinical outcomes**

S2.1. Clinical stimulation volume similarity to ‘gait-optimized’ stimulation volumes relates to gait outcomes

Compared to patients with gait improvement after DBS, patients whose gait worsened one year after DBS had clinical stimulation volumes which were significantly less similar to ‘gait-optimized’ stimulation volumes for the gait network (t = 3.60, p = 0.0006) (Supplementary Figure 8A). This pattern held when examining ‘gait-optimized’ stimulation volumes generated using the gait fibers (Supplementary Figure 8B), where patients with gait decline showed even larger deviations from their ‘gait-optimized’ stimulation volumes (t = 6.96, p < 0.0001).

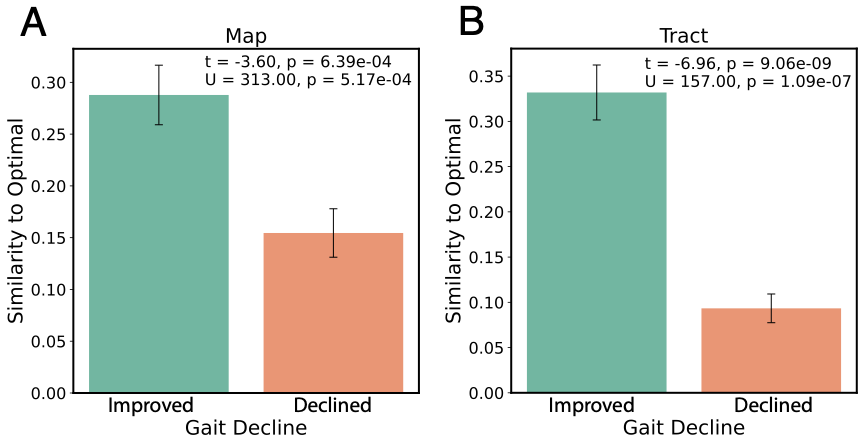

**Supplementary Figure 8. Worsening gait one year after DBS is associated with stimulation volumes far from theoretically optimal settings.** A) Patients with gait performance improvement versus gait performance decline one year after DBS. Patients with gait decline one year after DBS have significantly lower similarity to theoretically optimal stimulation volumes (t = 3.60, p = 0.0006). B) Similarity to tract-based gait target for patients with gait performance improvement versus gait performance decline one year after DBS. Patients with gait decline one year after DBS have significantly lower similarity to theoretically optimal stimulation volumes (t = 6.96, p < 0.0001).

S2.2. Similarity to ‘gait-optimized’ stimulation volumes relates specifically to gait outcomes

We next evaluated if similarity between the clinical stimulation volumes and ‘gait-optimized’ stimulation volumes was specifically associated with gait outcomes, or if it was a nonspecific effect related to other symptoms. Similarity between the clinician and ‘gait-optimized’ stimulation volumes did not correlate with changes in UPDRS-III total score or with tremor, rigidity, or bradykinesia subscores (Supplementary Table 1).

**Supplementary Table 1: Specificity of gait-optimized targets relative to other MDS-UPDRS III domains.** Correlations were tested between overlap with gait-optimized stimulation settings (functional or tract-based) and other MDS-UPDRS III subscores (total, tremor, rigidity, bradykinesia). No significant associations were observed, supporting the specificity of gait-optimized targets.

| **Optimization Target** | **Correlated MDS-UPDRS III Scores** | **Spearman Correlation** |
| --- | --- | --- |
| Gait (functional) | UPDRS-III Total | R = -0.14, p = 0.25 |
| Gait (functional) | Tremor | R = -0.12, p = 0.44 |
| Gait (functional) | Rigidity | R = 0.04, p = 0.76 |
| Gait (functional) | Bradykinesia | R = -0.07, p = 0.56 |
| Gait (fibers) | UPDRS-III Total | R = -0.07, p = 0.52 |
| Gait (fibers) | Tremor | R = -0.06, p = 0.70 |
| Gait (fibers) | Rigidity | R = 0.16, p = 0.22 |
| Gait (fibers) | Bradykinesia | R = -0.03, p = 0.81 |

S2.3. Optimal stimulation parameters for networks and fibers diverge

We wondered if the algorithm-suggested settings for network and fiber targets were converging on similar DBS parameters. Using the ‘optimized’ settings for the gait network and fibers, we found the two stimulation volumes diverged eve within the same patient (Supplementary Figure 9A). The one-sample t-test of Dice scores (overlap between network and fiber optimized stimulation volumes for each patient), was significantly below the cutoff threshold for ‘moderate’ similarity (Dice = 0.37±0.43) (t = 12.56, p < 0.0001), a Dice similarity coefficient of 0.6 (Supplementary Figure 9B).

We next investigated the difference between the stimulation volumes from the network- and tract-based targets. We compared the stimulation locations across the functional and tract-based targets within each patient and found the difference in stimulation parameters was driven by vertical displacement (t = 3.69, p_fwe_ = 0.0008) (Supplementary Figure 9C), primarily driven by asymmetry in the fiber-based targets with the right side being driven dorsally (3.10, p = 0.002) (Supplementary Figure 6).

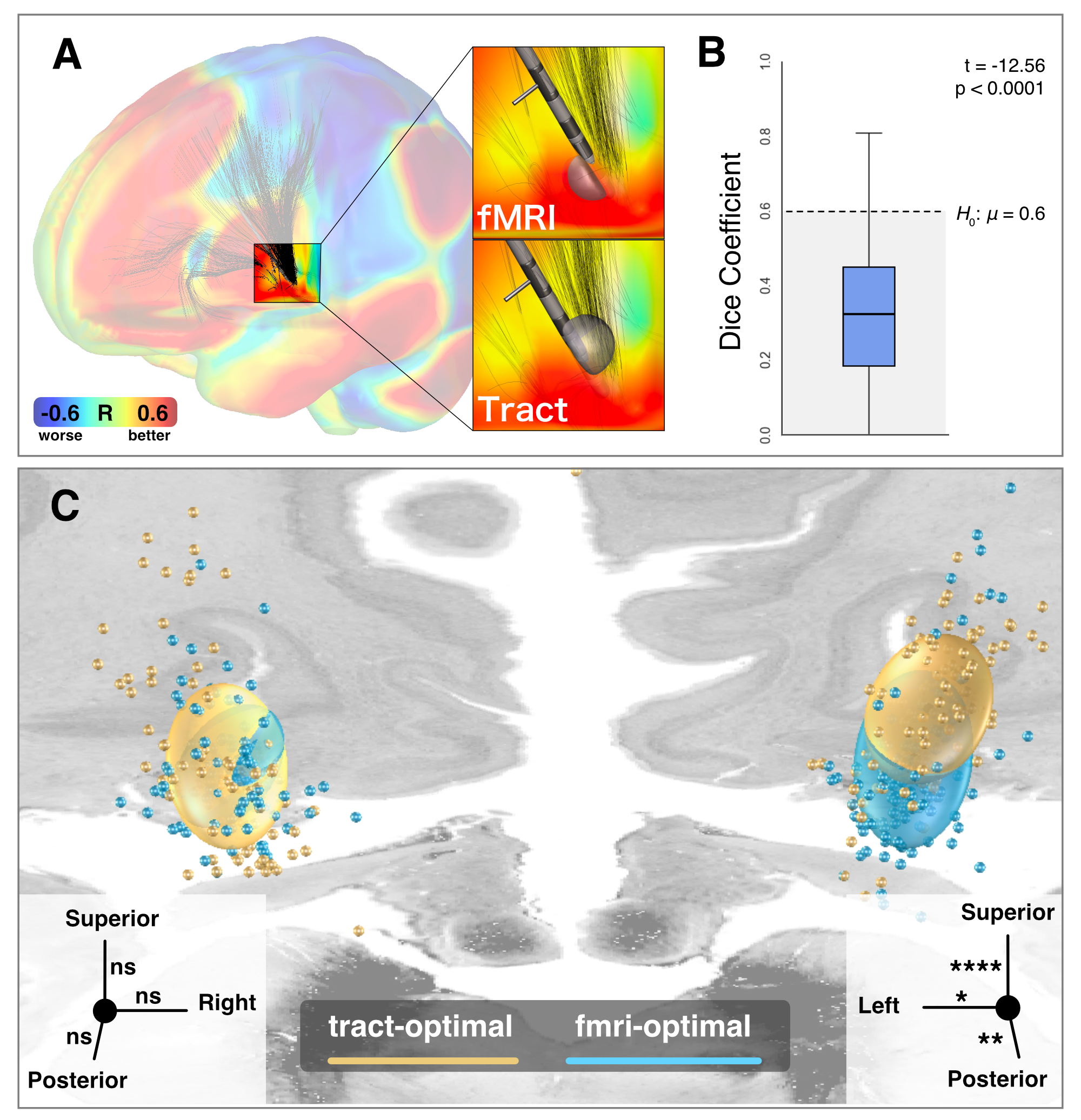

**Supplementary Figure 9. The gait network and fibers result in different DBS programs.** A) Example patient’s DBS electrode overlaid on the gait network and fibers, with the difference between the network-optimized and fiber-optimized stimulation volumes for this patient. B) The Dice coefficients of network- and fiber-based stimulation targets deviate significantly from achieving ‘moderate’ overlap at 0.6 (one sample t-test, t=12.56, p < 0.0001).^34^ C) The center coordinates of the network- and fiber-optimized stimulation volumes across the test cohort, with the ellipsoids representing the standard deviation of their axes. The fMRI-optimized centroids deviated from tracts centroids on the right (3.10, p = 0.002) but not the left (t = 1.2, p = 0.218). Individual deviations in the x, y, or z coordinates are shown in the axis inset. All p-values are FWE-corrected.

Programming to the functional gait target reduced stimulation of the tract-based target (t = 5.66, p < 0.0001), but did not result in overlap of stimulation volumes with negative tract regions (Supplementary Figure 10A). However, optimizing to the fiber-based target reduced overlap with the network-based target and included overlap of stimulation volumes with negative regions that could be associated with gait worsening (t = 12.31, p < 0.0001) (Supplementary Figure 10B).

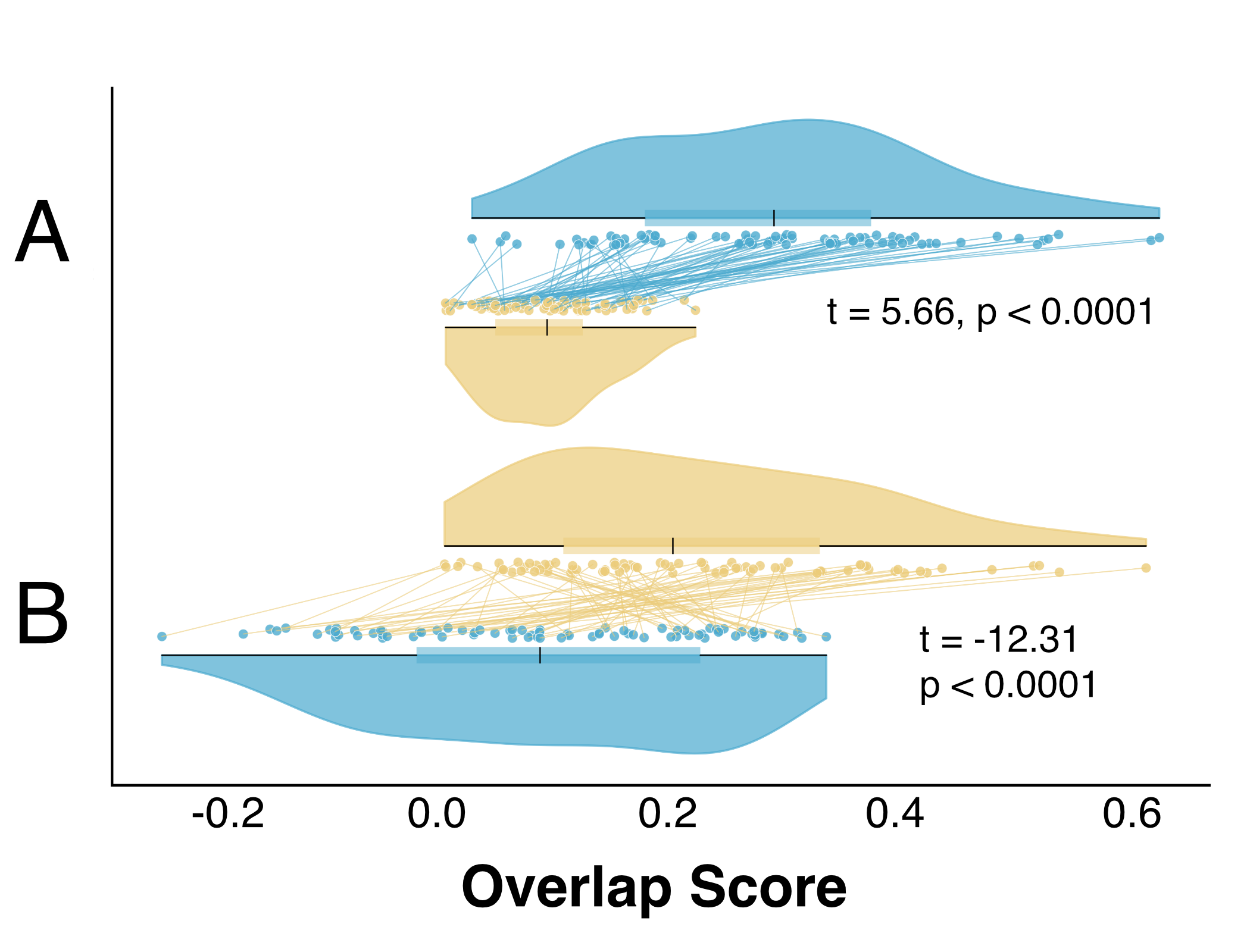

**Supplementary Figure 10. Optimizing to stimulate one gait-specific target compromises stimulation of the other target.** A) Stimulation volumes maximized to the gait network (blue) have significantly lower overlap with the gait fibers (yellow) (t = 5.66, p < 0.0001). B) Stimulation volumes maximized to the gait fibers have significantly lower overlap with the gait network (t = 12.31, p < 0.0001).

S2.4. Optimal gait outcomes are a joint function of increased similarity to both network- and fiber-based ‘gait-optimized’ stimulation volumes

We next investigated whether gait outcomes might be a function of increased overlap with ‘gait-optimized’ stimulation volumes from both targets. We derived two variables, one a measure of similarity between the patient’s clinical stimulation volume and the gait-network maximized stimulation volumes, and the other a measurement of similarity between the patient’s clinical stimulation volume and the gait-fiber maximized stimulation volumes.

We estimated the correlation of clinical and ‘maximized’ stimulation volume similarity derived from either target and measured their difference. This was repeated with 1,000 bootstraps to measure the reliability of the difference between these two targets. Tract-based targets showed stronger associations with clinical outcomes (t = 40.76, p < 0.0001), outperforming functional targets in 96% of simulations with an average correlation difference of 15% (ΔR = 0.15, p = 0.039) (Supplementary Figure 11). However, while the gait-fibers were consistently superior, they only outperformed the gait network by an average of 2% explained variance. Moreover, this effect was specific to gait, as there was no significant difference between the symptom-specific networks and fibers (Supplementary Figure 12) for the UPDRS (p = 0.21), tremor (p = 0.16), rigidity (0.20), and bradykinesia (p = 0.49).

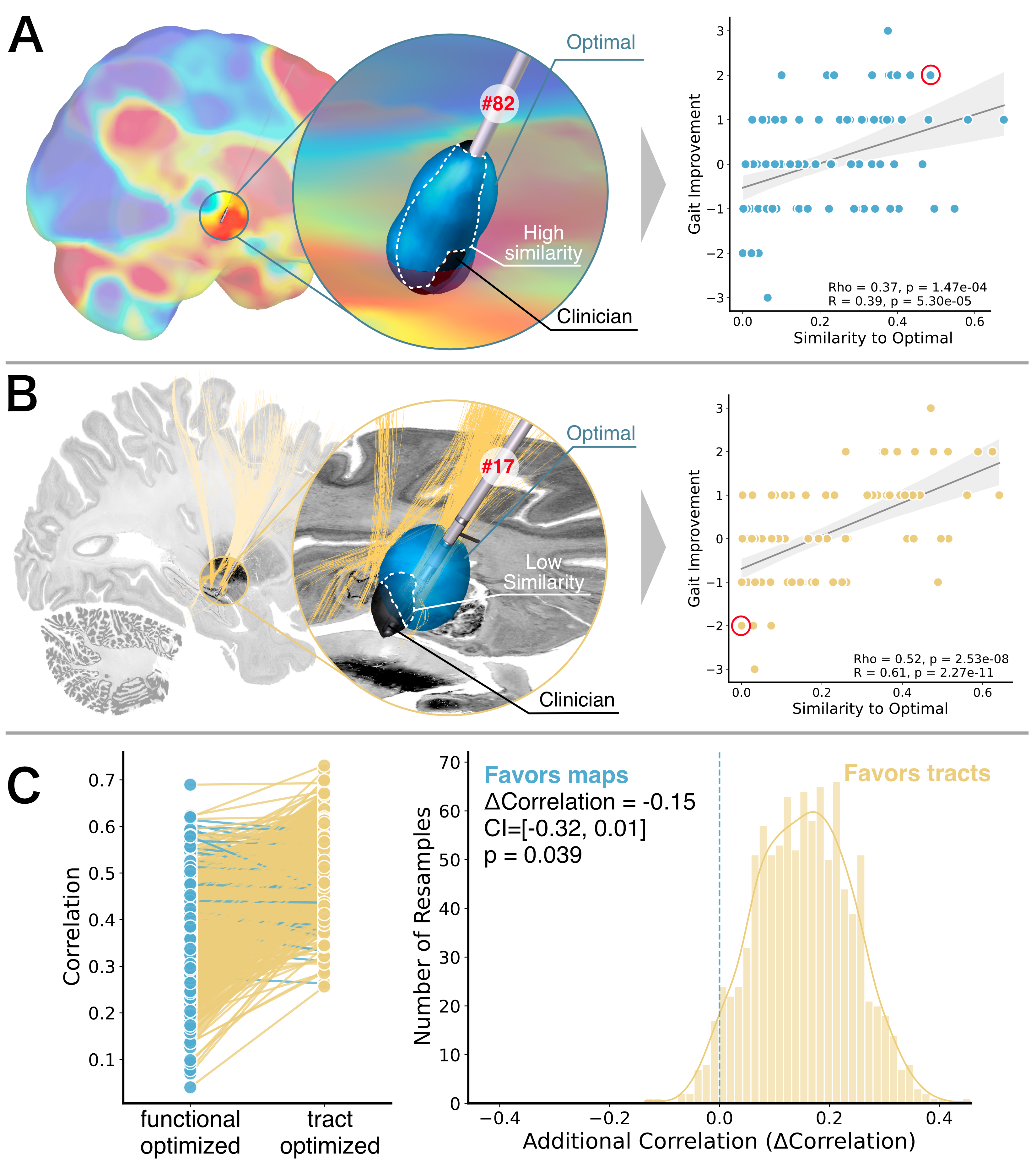

**Supplementary Figure 11.** Left) Pairwise bootstrapped correlations demonstrate the tract-based gait target has significantly stronger correlations with gait outcomes than the functional-based gait target (t = 40.76, p < 0.0001). Right) Superiority plot demonstrates similarity to fiber-based stimulation volumes is more strongly associated with gait outcomes than similarity to gait network stimulation volumes. This superiority is achieved 96% of the time, but explains, on average, 2% more variance (p = 0.0390).

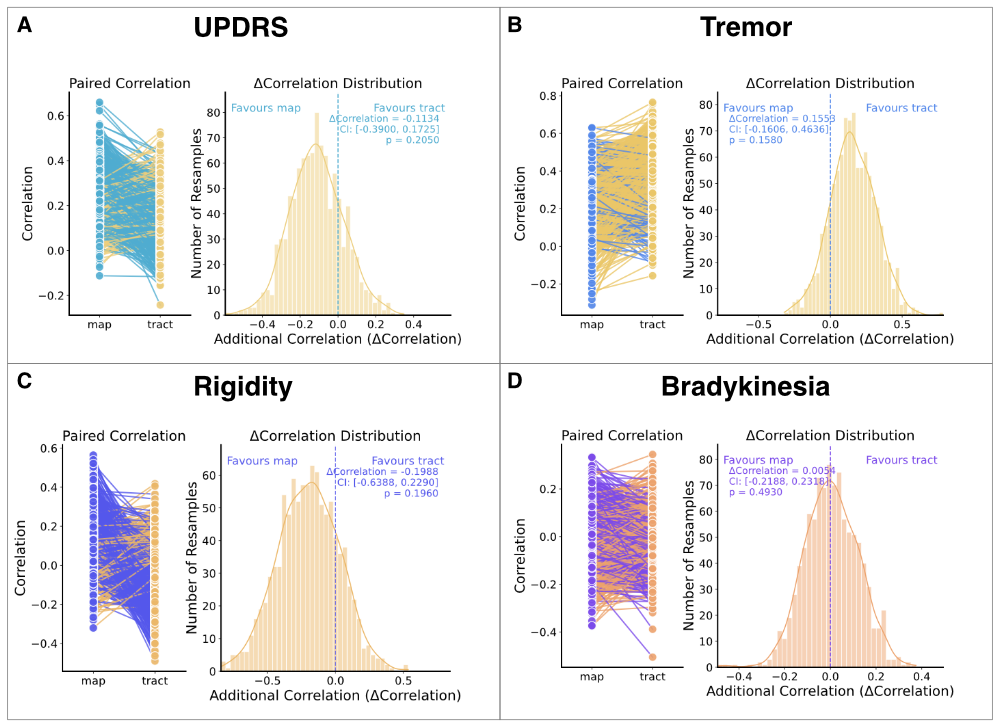

**Supplementary Figure 12. Similarity of clinical and symptom-specific stimulation volumes do not vary as a function of network or fibers.** A) Similarity of clinical and ‘UPDRS-optimized’ stimulation volumes do not have a significantly different relationship with UPDRS outcomes (p = 0.21). B) Similarity of clinical and ‘tremor-optimized’ stimulation volumes do not have a significantly different relationship with tremor outcomes (p = 0.16). C) Similarity of clinical and ‘rigidity-optimized’ stimulation volumes do not have a significantly different relationship with rigidity outcomes (p = 0.20). D) Similarity of clinical and ‘bradykinesia-optimized’ stimulation volumes do not have a significantly different relationship with bradykinesia outcomes (p = 0.49). Left panels show paired correlation values between functional- and tract-based targets for individual patients. Right panels show distributions of correlation differences (Δ Correlation) from resampling analyses, with confidence intervals and p-values shown in each panel.

S2.5 Optimal gait outcomes are a joint function of increased similarity to both network- and fiber-based ‘gait-optimized’ stimulation volumes

While the fibers outperformed the network in univariate analyses, we investigated how these related to gait outcomes in a multivariate analysis. When these two variables were combined in a linear model regressing on gait outcomes, increased overlap with network-based stimulation volumes was significantly related to outcomes (t = 3.10, p = 0.0029), although similarity to the fiber-based stimulation volumes was not (t = -0.38, p = 0.71). However, they did interact such that the patients with the best outcomes were those with high overlap with both targets (t = 4.08, p = 0.0001). The lack of individual significance in the fiber-based target is likely due to the significant collinearity with functional-based targets (r = 0.74, p < 0.0001), as well as inherent collinearity with the interaction term. This specifically suggests that network- and fiber-based stimulation volumes have similar amounts of overlap with their respective clinical stimulation volumes, indicating that the stimulation volumes derived from either target are partially similar.

S2.6. Relationship between outcomes and optimal stimulation programs for other PD symptom domains

We next evaluated if the association of clinical outcomes with the similarity between clinical and ‘symptom-optimized’ stimulation volumes might also relate to outcomes in other symptoms. To do this, we generated ‘symptom-optimized’ stimulation volumes using networks and fibers for the UPDRS-III, tremor, rigidity, and bradykinesia. For both networks and fibers, similarity between clinical and ‘symptom-optimized’ stimulation volumes was significantly associated with post-operative outcomes for the UPDRS (average r = 0.25, p = 0.028) and tremor (average r = 0.30, p = 0.045), but not bradykinesia (average r = 0.11, p = 0.49) nor rigidity (average r = 0.08, p = 0.57) (Supplementary Figure 13).

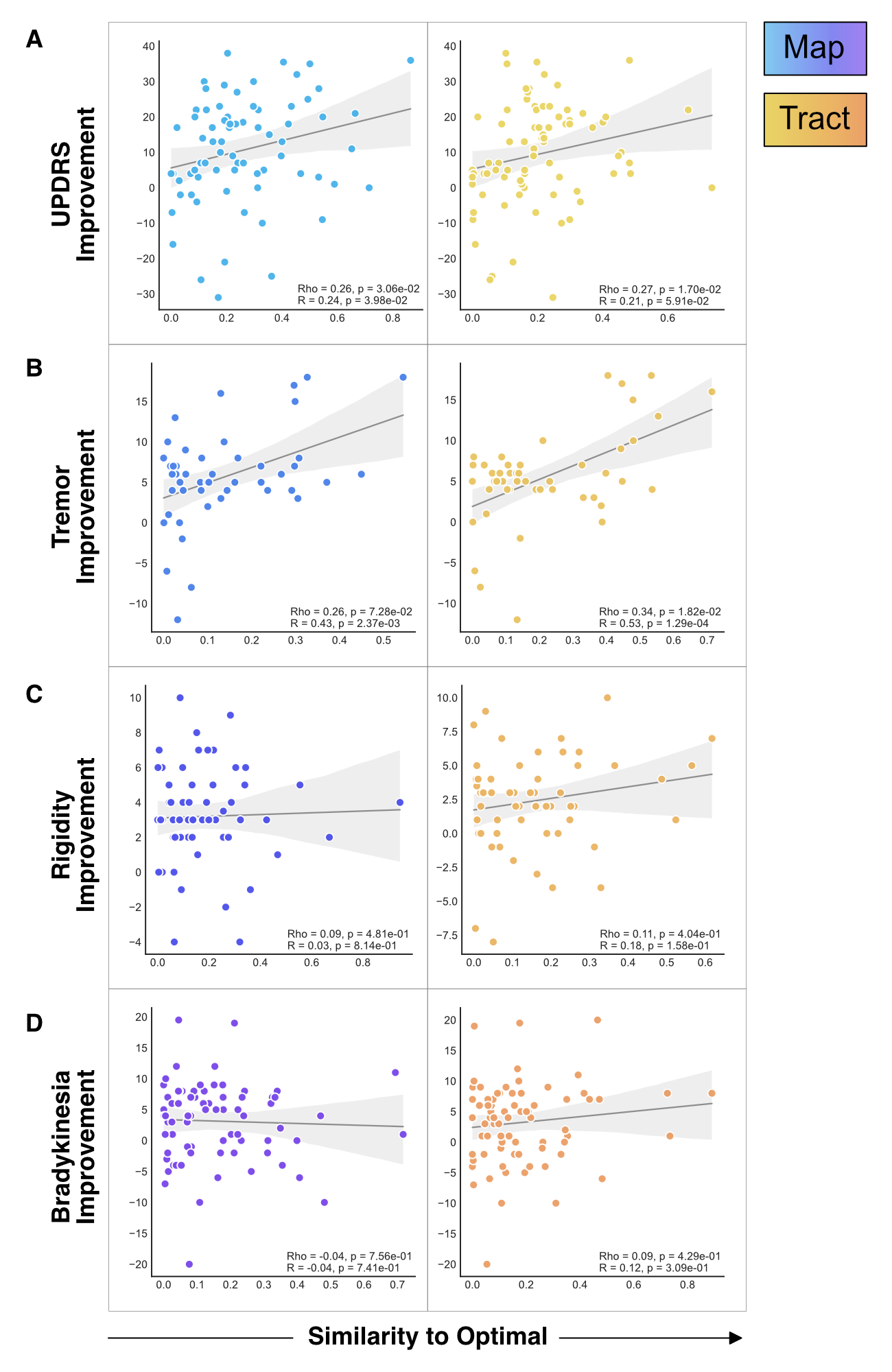

**Supplementary Figure 13. Similarity of clinical and ‘symptom-optimized’ stimulation volumes are related to outcomes in some symptoms.** A) Similarity of clinical and ‘UPDRS-optimized’ stimulation volumes derived from the UPDRS network (left) and fibers (right) are associated with UPDRS outcomes after DBS (average r = 0.25, p = 0.028). B) Similarity of clinical and ‘tremor-optimized’ stimulation volumes derived from the tremor network (left) and fibers (right) are associated with tremor outcomes after DBS (average r = 0.30, p = 0.045). C) Similarity of clinical and ‘rigidity-optimized’ stimulation volumes derived from the rigidity network (left) and fibers (right) are not associated with rigidity outcomes after DBS (average r = 0.11, p = 0.49). D) Similarity of clinical and ‘bradykinesia-optimized’ stimulation volumes derived from the bradykinesia network (left) and fibers (right) are not associated with bradykinesia outcomes after DBS (average r = 0.08, p = 0.57).

S2.7. Similarity to ‘symptom-optimized’ stimulation volumes explains variance differentially

We next investigated if similarity between the clinical and ‘gait-optimized’ stimulation volumes explained more variance in gait outcomes than other ‘symptom-optimized’ stimulation volumes could explain in their respective symptoms (tremor, rigidity, bradykinesia, and overall UPDRS-III). To do this, we computed the Spearman correlation between outcome in each symptom’s outcome and that symptom’s clinical-‘symptom-optimized’ stimulation volume similarity. Then, we measured the explained variance (ρ^2^) in each symptom and calculated the pairwise differences. To measure significance of these differences, we permuted the data and repeated the correlations 1000 times to derive a null distribution. Similarity of clinical to ‘gait-optimized’ stimulation volumes explained, on average, 22% more variance than other ‘symptom-optimized’ stimulation volumes did for their respective symptoms (all p < 0.0001) (Supplementary Figure 14), represented by the left column of the two heatmaps. We want to emphasize this experiment represents the difference in variance explained between ‘symptom-optimized’ stimulation volumes and their own symptoms (i.e. ‘gait-optimized’ stimulation volume and gait outcomes), not the ability for cross-explanation of one ‘symptom-optimized’ stimulation volume with another symptom (i.e. ‘gait-optimized’ stimulation volumes and rigidity outcomes).

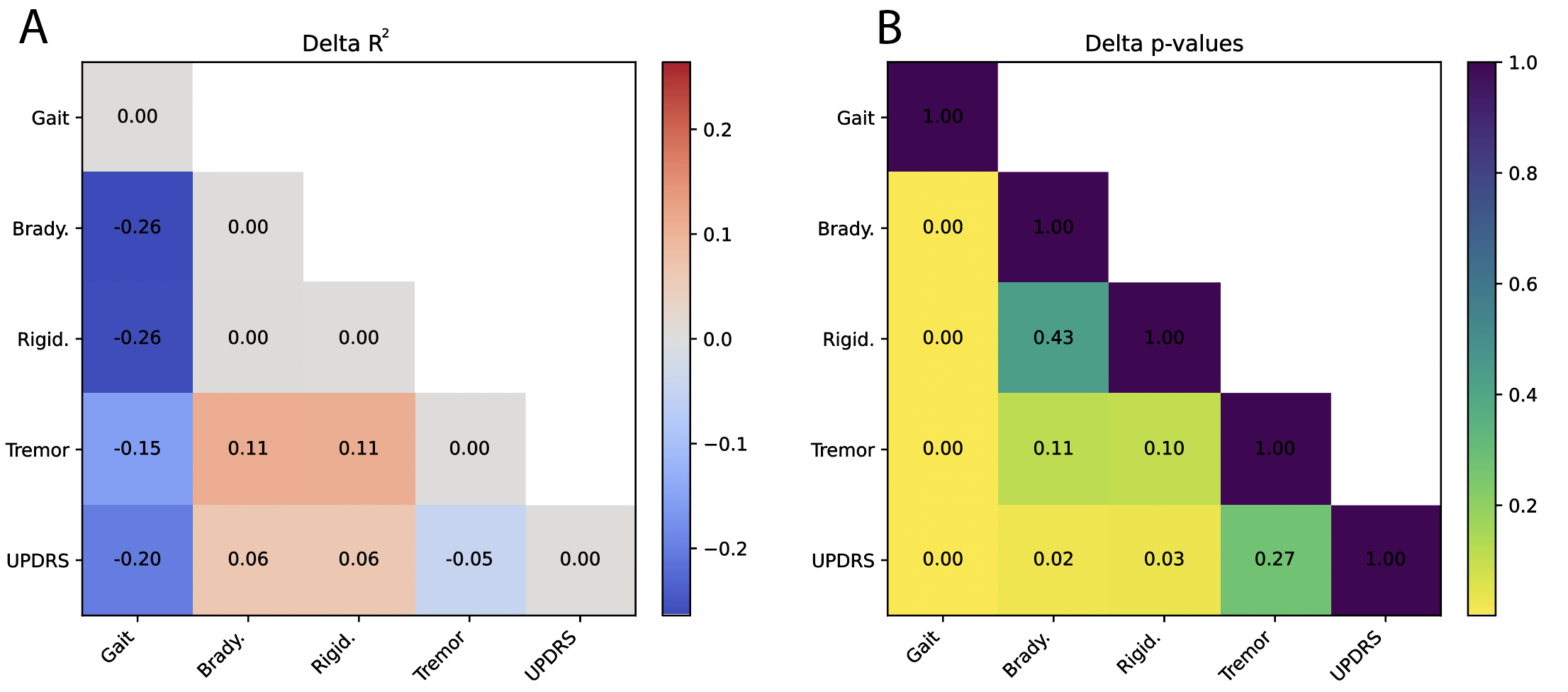

Supplementary Figure 14. Difference in explained variance of symptoms by similarity of clinical stimulation volumes to their respective symptom-specific stimulation volumes. A) ∆ρ^2^ measured across all comparisons. B) Permutation-based p-values from associated ∆ρ^2^ comparisons. Squares within the heatmap represent pairwise comparisons of explained variance, comparing the explained variance of the two symptoms by similarity of their clinical and ‘symptom-optimized’ stimulation volumes. This represents the average explained variance across the ‘symptom-optimized’ stimulation volumes generated by targeting networks and fibers within a symptom.

**Supplementary Materials 3: Prospective analysis**

S3.1 Detailed clinical features of the prospective cohort

In the prospective reprogramming cohort, we investigated the specific clinical factors in each patient, including their pre-surgical gait score (Supplementary Table 2).

**Supplementary Table 2: Clinical characteristics of prospectively reprogrammed cohort.** Patients were selected based on gait impairment as the chief clinical complaint and evaluated in the OFF-medication state. Carbidopa-levodopa regimens are reported with respect to dosage (CR: controlled-release). Pre-surgical gait severity is reported using item ‘3.10: Gait’ from the MDS-UPDRS III clinical scale.

| **ID** | **Age** | **Years from DBS** | **Medication State** | **Medication Routine**  **(carbidopa-levodopa mg)** | **Pre-surgical gait severity (MDS-UPDRS III Item 3.10)** | **Chief Complaint** |
| --- | --- | --- | --- | --- | --- | --- |
| 1 | 50-60 | 1 | OFF | 25-100 mg + CR | 2 | Gait |
| 2 | 50-60 | 1 | OFF | 25-100 mg | 3 | Gait |
| 3 | 60-70 | 3 | OFF | 25-100 mg | 2 | Gait |
| 4 | 60-70 | 2 | OFF | 25-100 mg + CR | 2 | Gait |
| 5 | 70-80 | 6 | OFF | 25-100 mg | 4 | Gait |
| 6 (unilateral) | 60-70 | 3 | OFF | 25-100 mg | 2 | Gait |

S3.2. Prospective cohort ‘gait-optimized’ DBS settings

Consistent with our retrospective analyses, we assessed whether the ‘gait-optimized’ settings were associated with increased overlap of the gait target compared to each patient’s clinical settings. We should emphasize this analysis was performed post-hoc as a verification analysis. We normalized each patient’s overlap to their own clinical setting to generate a measurement of increased gait target overlap relative to baseline. Under this within-patient framework, the gait-optimized settings demonstrated a significantly higher overlap with the gait target compared to clinical settings (t = 2.9, p = 0.016), indicating that the algorithm successfully increased stimulation of the intended target under real-world programming conditions (Supplementary Figure 15).

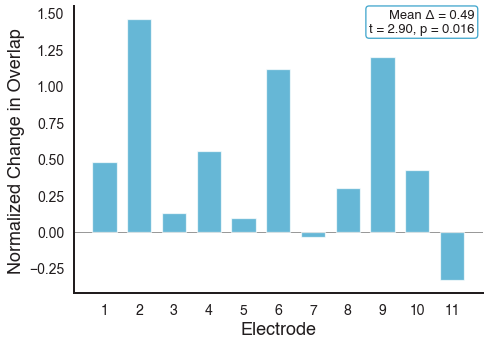

**Supplementary Figure 15. Per-electrode overlap of ‘gait-optimized’ stimulation volumes with the chosen gait target in the prospective patients.** Bar plot showing the normalized change in overlap with the gait targets for each electrode in the prospective cohort, comparing ‘gait-optimized’ settings to each patient’s clinical settings. Zero represents baseline overlap of the clinical stimulation volumes with each patient’s chosen gait target (network or fibers). Positive values indicate greater overlap with the gait target under gait-optimized programming relative to clinical programming. Across patients, gait-optimized stimulation volumes had a significantly higher overlap with the chosen gait target (t = 2.90, p = 0.016).

**Supplementary Materials 4: Optimization Algorithm Details**

S4.1. Fast geometric approximation of stimulation volumes to enable iteration

The core of our optimization algorithm uses fast approximation of stimulation volumes to enable a full algorithmic convergence within several seconds on standard clinical computational infrastructure. While prior algorithms use precise biophysical modelling, we reformulate this as a geometry problem where we need to solve the location, size, shape, pitch, roll, and yaw of stimulation volumes. Using the electrode localization and model information from Lead-DBS,^30^ we extract each contact’s coordinate and use this to begin our geometric approximation of the stimulation volume (**Figure 1B**).

We first discuss the simplest approximation, which is the stimulation volume at undirected contacts. We formulate the stimulation as a sphere defined by all points within the radius of the stimulation volume. The radius of the stimulation volume can be defined as $r(a)=\sqrt{(\frac{a-0.1}{0.22})}$, where $a$ is the current amplitude provided to that contact in milliamps, these two constants having been previously derived empirically.^31^ Thus the stimulation volume at a contact is then $\left\| \boldsymbol{x}-\boldsymbol{c} \right\|_{\boldsymbol{2}}<r(a)$ where $\boldsymbol{x}$ is the coordinate vector being checked, and $\boldsymbol{c}$ is the central coordinate vector of the stimulation volume (the contact’s center).

We next discuss fast approximation of stimulation volumes at directional contacts. This is formulated as the stimulation which is both within the stimulation volume’s radius but not the components of the stimulation which would be eliminated by electrical shielding of the neighboring contacts. To account for electrical field shielding in directional contacts, we calculate the midpoint, $\boldsymbol{m}=\frac{1}{2}\boldsymbol{(c}_{1}+\boldsymbol{c}_{2})$ between contact coordinates pairs ($\boldsymbol{c}_{1}$and $\boldsymbol{c}_{2}$) as well as the connecting vector between the two points, $\boldsymbol{n}=\boldsymbol{c}_{1}- \boldsymbol{c}_{2}$. The midpoint and connecting vector can be used to generate an orthogonal plane between the two contacts, $\mathbf{P}_{1,2}=\boldsymbol{m}^{\boldsymbol{T}}\boldsymbol{n}+d$ where $d$ is an offset is solved for by setting $0=\boldsymbol{m}^{\boldsymbol{T}}\boldsymbol{n+}d$. Any point on this plane is zero-valued, while points behind it (closer to $\boldsymbol{c}_{\boldsymbol{2}}$) are negative and points in front of it (closer to $\boldsymbol{c}_{\boldsymbol{1}}$) are positive. This allows us to define our directional stimulation volume as $\boldsymbol{x}^{\boldsymbol{T}}\boldsymbol{n}_{\boldsymbol{1,2}}\boldsymbol{+}d\boldsymbol{>}0$, where $\boldsymbol{x}$ is the coordinate of interest and $\boldsymbol{n}_{\boldsymbol{1,2}}$ is the vector connecting the two contacts. Coordinates that are both within the radius of the stimulation volume and that pass this test for all neighboring contacts are within the directional stimulation volume.

The overall stimulation volume can be represented as the union of each individual stimulation volume by $\mathbf{V}=\bigcup_{i=1}^{N} \mathbf{V}_{i}$ where each $\mathbf{V}_{i}$ is the stimulation volume at contact $i$ (and $i = 1 . ..N$) where $N$ is the total number of contacts (**Figure 1C**).

S4.2. Converting functional- and tract-based symptom-specific targets into optimization targets

The ideal stimulation settings to maximally stimulate a connectomic target can be formulated as a non-convex optimization problem which must be solved for each patient and electrode individually. The goal of this algorithm maximizes the density of positive (beneficial) targets within the overall stimulation volume while minimizing the density of negative (deleterious) targets within the overall stimulation volume. To optimize to network targets, we measured the connectivity of each brain voxel to the network of interest,^7^ generating a description of how connectivity to that symptom-specific target varies across the brain.^32^ This ‘symptom target map’ represents the optimization target. To optimize to tract-based targets, we use symptom-specific fibers and calculate their tract densities,^16^ converting them to voxelwise formats. This defines a description of how connectivity to those symptom-specific fibers varies across space.^8^ This results in a common optimization target format for functional- and tract-based targets.

S4.3. Optimization of stimulation volumes to symptom-specific connectomic targets

We next developed a loss function which would increase the positive values within the stimulation volume while avoiding dangerous amounts of current or indiscriminate expansion of stimulation across the brain. First, we define a target function to maximize the density of positive values (from either functional- or tract-based targets) within its stimulation volume as $G\left( \boldsymbol{V} \right)=\frac{\int_{\boldsymbol{V}} \boldsymbol{T} d\boldsymbol{V}}{\left| \boldsymbol{V} \right|}$, where $\boldsymbol{V}$ is the stimulation volume and $\boldsymbol{T}$ is the functional- or tract-based target.

Potentially dangerous levels of amperage can be avoided by a penalty function, $F(\boldsymbol{a)=}\frac{\lambda}{b\boldsymbol{-}min\boldsymbol{(}b\boldsymbol{,}\sum_{i\boldsymbol{=}1}^{N} {max\boldsymbol{(}0,\boldsymbol{a}}_{i}\boldsymbol{))}}$, where $b$ is a scalar blocking constant representing the amplitude that cannot be exceeded and $\lambda$ is a hyperparameter which scales the penalty and $i$ is the contact index (and $i = 1 . ..N$) with $N$ being the total number of contacts. As the sum of positive amperages (active contacts) in $\boldsymbol{a}$ approaches the blocking value, $b$, the penalty function approaches infinity. We set $b=5$ given the common clinical practice to avoid passing over 5 milliamps across any given electrode.^33^

Further, a small penalty can gently penalize each contact to avoid passing current unless it provides non-negligible benefits with $\mathcal{H}\left( \boldsymbol{a} \right)\boldsymbol{=}\gamma\sum_{i\boldsymbol{=}1}^{N} {max(0\boldsymbol{, a}}_{i}\boldsymbol{)}$,

where $\gamma$ is again a hyperparameter which scales the penalty and $i$ is the contact index (and $i = 1 . ..N$) with $N$ being the total number of contacts. The final merit function then becomes $\mathcal{L}\left( \boldsymbol{V,a} \right)\boldsymbol{=}G\left( \boldsymbol{V} \right)$ – $F$($\boldsymbol{a}$) – $\mathcal{H}$($\boldsymbol{a}$). This merit function can then be maximized using gradient-based techniques. Here we perform gradient ascent with gradient clipped adaptive moment estimation (**Figure 1C**).^34^

S4.4. Hyperparameter tuning

We tuned the hyperparameters ($\gamma and \lambda$) using a grid search in the training cohort. The optimization was run with each set of hyperparameters (varied across values of 0.001, 0.01, 0.1, 1, 10) and the final loss was recorded across all searches. This process was repeated for both functional- and tract-based targets. Then, the optimal hyperparameters were averaged to identify the hyperparameter for both.

S4.5. The initial guess for stimulation volume optimization

Optimization problems and especially non-convex problems are sensitive to the initial guess. In our case, the initial guess problem depends upon the initial amplitude vector passed to the optimizer. We developed an initial guess procedure which uses triangulation to identify an approximation of the amplitude to apply across the contacts.

For each contact with coordinate $c_{i}$, we start by defining a field around the contact defined by a sphere of radius $r=2.55 mm$. Within this field, we identify local maxima of the network- or tract-based target, which correspond to spatial positions where stimulation is expected to be most beneficial. For each local maximum, we extract its centroid $m_{j}$ and magnitude $s_{j}$. We then quantify the distance $d_{ij}$ between the contact coordinate *i* and the local maximum centroid *j* using the Euclidean distance. Contacts that are closer to maxima with high magnitude are preferentially weighted using a soft proximity kernel that decays smoothly with distance. This weighting allows us to assign each contact a score defined as $r_{i}= \sum\frac{s_{j}}{1+exp(d_{ij}- 1)}$. These contact relevance scores can then be normalized across contacts to obtain weights which are used to allocate an initial current distribution to each of the contacts.

This triangulated initialization defines the starting amplitude vector for the full optimization of the stimulation volume but does not constrain the final solution, which is determined by maximizing the loss function through gradient ascent. To evaluate convergence properties, we compared this weighted initialization to a Gaussian-distributed initial guess by re-running the optimization 100 times per patient and measuring the mean and standard deviation of the resulting stimulation volumes. We additionally compared the achieved loss values across initialization strategies in the training dataset, demonstrating improved stability and performance with the triangulated approach.

S4.6. Projection of optimized parameters

The optimization algorithm maximizes the density of a given target within the stimulation volume. This minimizes the current required to intersect a target, but can result in very small current volumes around local maxima. Thus, the optimization algorithm may paradoxically apply small currents to the ideal contacts which may often be below meaningful stimulation thresholds. Thus, we developed a final step which projects the currents across the electrode’s contacts into clinically realistic amplitudes, prioritizing allocation of current to contacts near local maxima.

To do this projection, we iterate over each contact, $i$, across the total number of contacts, $N$, where $i = 1 . ..N$. For each contact, we calculate its merit using the merit function defined above (Supplementary Materials 4.3), $\mathcal{L}\left( \mathbf{V}_{i}\boldsymbol{,}\boldsymbol{a}_{i} \right)\boldsymbol{,}$ where $\mathbf{V}_{i}$ is the stimulation volume at that contact and $\boldsymbol{a}_{i}$ is the current amplitude. We then normalize this across the merit of all electrodes, $\sum_{i}^{N} \mathcal{L}\left( \mathbf{V}_{i}\boldsymbol{,}\boldsymbol{a}_{i} \right)$**,** and scale it by the total amplitude allocated across all contacts, $\sum_{i}^{N} \boldsymbol{a}_{i}$. This weighting allows us to assign each contact a score defined as $a_{i}= \sum_{i}^{N} \boldsymbol{a}_{i}\frac{\mathcal{L}\left( \mathbf{V}_{i}\boldsymbol{,}\boldsymbol{a}_{i} \right)}{\sum_{i}^{N} \mathcal{L}\left( \mathbf{V}_{i}\boldsymbol{,}\boldsymbol{a}_{i} \right)}$. This reallocates the overall amplitude, prioritizing contacts near strongly positive regions. A limitation of this projection is that it may reduce merit, and thus overlap, with the target network. However, it results in increasing amperage at the strongest contacts to meaningful values.
